# Epidural Versus Transcutaneous Spinal Cord Stimulation For Motor Recovery After Spinal Cord Injury: A Comparative Analysis

**DOI:** 10.64898/2026.06.22.26356277

**Authors:** Shovan Bhatia, Roberto M. de Freitas, John H. Kanter, Thomas J. Buell, David O. Okonkwo, Elvira Pirondini, Genís Prat-Ortega, Marco Capogrosso, Peter C. Gerszten

## Abstract

Spinal cord injury (SCI) is a devastating neurological injury that results in the profound loss of voluntary motor function and marked reduction in quality of life. Rehabilitation remains as the standard of care for recovery after SCI; however, it often falls short in recovering meaningful motor function. Spinal cord stimulation (SCS) has emerged as a promising neurostimulation approach to fill this gap and recover lost voluntary motor function. Two main approaches of SCS have been designed and implemented for human use: epidural and transcutaneous SCS. Over the last two decades, several clinical studies have shown convincing evidence that both epidural and transcutaneous SCS can be used in conjunction with rehabilitation to improve motor function of individuals after SCI. Yet fundamental clinical questions remain unanswered: when should clinicians choose epidural or transcutaneous SCS, which technique provides the most durable outcomes, and for whom is each therapy best? Without these answers, widespread and meaningful adoption of either approach into clinical practice will remain limited. To address these questions, in this Review, we define the distinct therapeutic goals, intended use cases, clinical parameters, and responder profiles for both epidural and transcutaneous SCS to guide their eventual adoption into clinical practice. We found that indeed epidural and transcutaneous SCS serve distinct therapeutic roles. Epidural SCS is designed as an assistive therapy that can restore muscle activity and single joint movements immediately within one week of implantation, while transcutaneous SCS is designed as a long-term therapeutic device with cumulative functional gains observed over treatment periods of up to 18 weeks. Lastly, epidural SCS produced benefits for all participants (AIS A-D) despite the extent of their injury, while transcutaneous SCS only consistently benefits individuals with incomplete motor injuries (AIS C-D).

## INTRODUCTION

### Spinal cord injury

Traumatic spinal cord injury (SCI) is a devastating neurological injury that results in severe paralysis and affects over 300,000 people living with SCI in the United States^1–3^. SCI survivors often are left with a profound loss of functional independence and medical sequelae that severely impacts their quality of life^1^. Notably, less than 1% of patients are discharged from the hospital with complete recovery of their paralysis^2^. Thus, diligent participation in rehabilitation regimens is critical to give patients the best chance at recovering motor function. Conventional rehabilitation strategies emphasize achieving independent mobilization and optimizing functional recovery in the at-home setting; however, these approaches largely fall short to recover meaningful motor function^4–6^.

Over the past two decades, neurostimulation has emerged as a compelling adjunct to conventional rehabilitation following SCI, designed to remediate the inefficacy of traditional rehabilitation protocols^7–10^. In particular, spinal cord stimulation (SCS) is one such promising approach that has a growing body of clinical evidence supporting its ability to restore lost motor function^11–16^. Two approaches of SCS have been developed for human use: epidural (eSCS) and transcutaneous SCS (tSCS). eSCS is considered an assistive neurostimulation device that immediately improves motor function as soon as stimulation is turned on, while tSCS is a therapeutic neurostimulation device that restores functional gains gradually over weeks of high-intensity rehabilitation. While evidence has grown to support both approaches, fundamental clinical questions are still left unanswered, limiting widespread adoption of either approach: (1) When should clinicians choose eSCS or tSCS? (2) What are the differences in the goals of each modality? (3) What are the ideal configurations that maximize benefit? (4) Who are the ideal responders to either technique? Before meaningful adoption of eSCS or tSCS can occur, these fundamental clinical questions must be addressed.

Thus, this Review was performed in order to systematically address the aforementioned questions and provide a timely and critical analysis of the contemporary literature evaluating epidural (eSCS) and transcutaneous SCS (tSCS) as key neurostimulation approaches for facilitating upper and lower limb motor recovery after SCI (**Figure 1**). We prioritized studies reporting individual *de novo* participant-level data to inform direct comparisons between eSCS and tSCS. Here, we define the distinct therapeutic goals, intended use cases, clinical parameters, and responder profiles for both eSCS and tSCS, and provide a framework to guide their rational and consistent adoption into clinical practice.

**Fig. 1.**
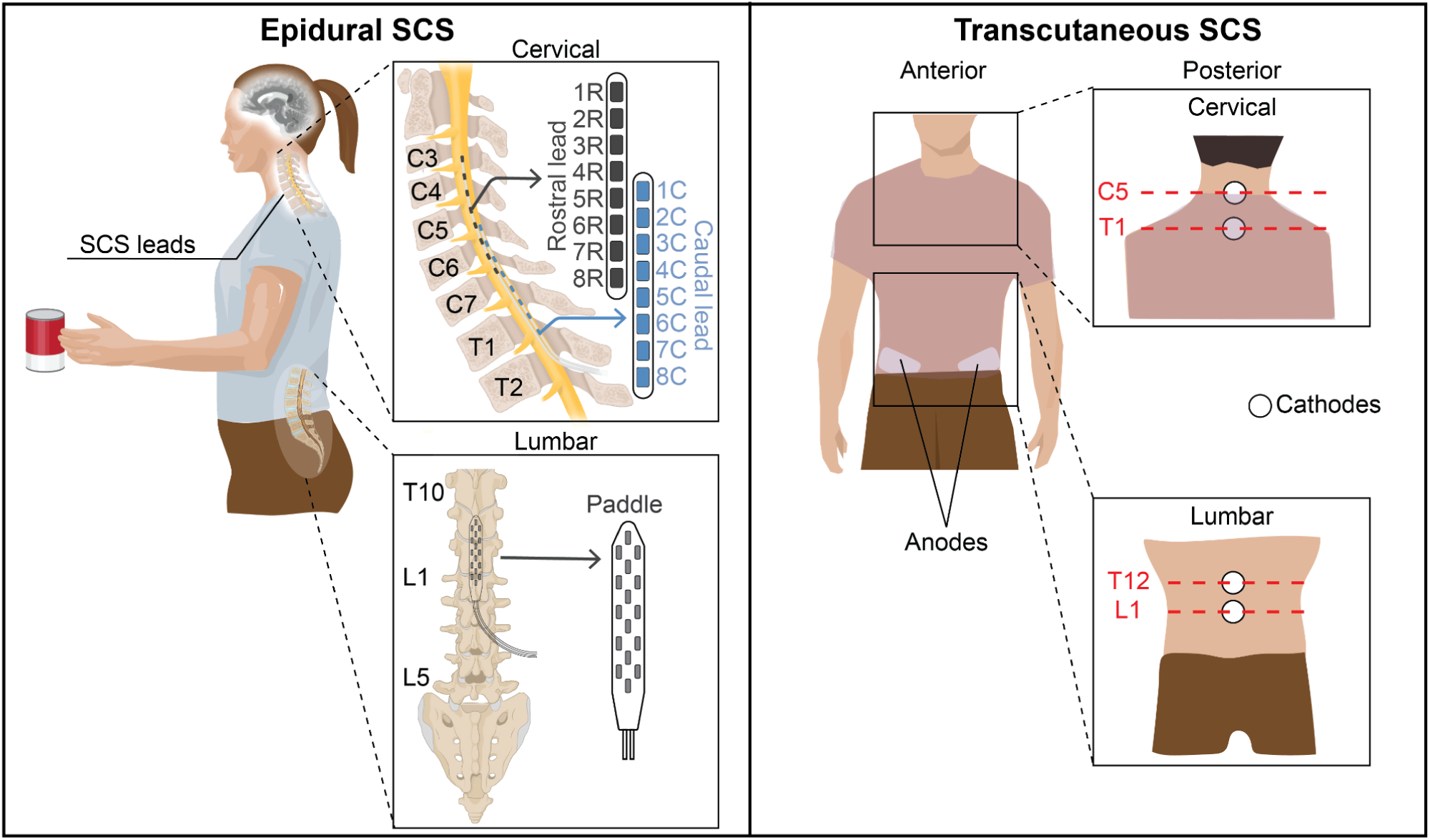
Overview of epidural and transcutaneous SCS. Epidural SCS are surgically implanted leads positioned within the epidural space. For upper limb applications, two linear percutaneous leads are arranged rostrocaudally spanning the cervical motor pools from C3-T1. For lower limb applications, a paddle lead is implanted over the lumbosacral enlargement (T10-L2). Transcutaneous SCS delivers stimulation non-invasively via surface electrodes. One or two adhesive cathodes placed over the posterior midline at the C5–T1 or T12–L1 vertebral levels, with return anodes positioned anteriorly over the abdomen or iliac crests.

### Spinal cord stimulation (SCS)

#### Epidural SCS

##### Overview

Epidural SCS is a neurotechnology that was first introduced in the late 1960s for treating patients with intractable pain^17^. By targeting the dorsal sensory roots with high-frequencies (often >1kHz), eSCS was able to effectively dampen the aberrant proprioceptive pain signals^17,18^. As this technology was slowly adopted into clinical practice, serendipitous improvements in voluntary motor control were observed in a patient with multiple sclerosis who was receiving eSCS therapy for her intractable pain^19^. Following this, several animal, human and computational studies were designed and carried out to understand the effects of eSCS for recovering voluntary function after paralysis^11–13,20,21^.

Implantation involves a surgical procedure in which epidural leads are placed using image guidance over the dorsal sensory afferents at spinal levels corresponding to the cervical or lumbosacral enlargement. Intraoperative neuromonitoring is often used to precisely position the leads. Typically, percutaneous leads are preferred for the cervical spinal cord, while paddle electrodes are preferred for the lumbar spinal cord due to the size of the spinal canal (**Figure 1**). Accurate lead positioning is critical for durable efficacy and long-term usability^22^. Intraoperative fluoroscopy and neurophysiological monitoring can be valuable adjuncts to guide accurate placement. Postoperative recovery is generally rapid, with most patients discharged on the day after surgery with return to normal daily activity within 1 week.

##### Mechanism of action

The mechanism by which eSCS improves motor function is well established. Epidural stimulation at low frequencies (40-60Hz) selectively recruits large Ia-afferent sensory fibers that synapse through mainly monosynaptic projections onto the α-motoneuron^23^. Importantly, there are two main stimulation paradigms of eSCS: continuous vs. spatiotemporal stimulation. Continuous eSCS delivers tonic stimulation that immediately facilitates voluntary movement by modulating the membrane potential of motoneurons to transform subthreshold excitatory postsynaptic potentials (EPSPs) into suprathreshold action potentials, enabling control of previously paralyzed limbs^24^. Spatiotemporal eSCS, by contrast, sequences stimulation across multiple electrode contacts to mimic the natural spatiotemporal activation patterns of the spinal cord during locomotion. However, this paradigm requires pairing with an external interface capable of decoding motor intent and translating it into SCS commands in real time^25^. For the purposes of this Review, continuous and spatiotemporal stimulation paradigms are considered collectively under eSCS.

##### Key clinical trials

Early studies by Dimitrijevic and colleagues showed that eSCS of the lumbosacral spinal cord in individuals with complete paraplegia could activate the locomotor central pattern generator (CPG), producing rhythmic, locomotor-like electromyography (EMG) activity and stepping movements even with non-patterned stimulation to facilitate ambulation, thereby laying the foundation for clinical trials of eSCS^26^. Three landmark studies demonstrated the use of SCS for patients with motor incomplete^13^ and motor complete paralysis^12,14^.

In 2011, Harkema et al. demonstrated that combining intensive rehabilitation with eSCS enabled a participant with AIS B SCI to regain voluntary standing and lower limb movements after 7 months. This established the critical role of load-bearing proprioceptive input and intrinsic spinal sensorimotor circuitry in functional recovery. However, in 2014, Angeli et al. expanded these findings to a larger cohort, including individuals with sensory complete AIS A injuries, demonstrating voluntary activation of previously paralyzed muscles and established evidence for broader clinical application. In 2018, Angeli et al. further showed that intensive locomotor training combined with eSCS restored voluntary EMG activity and single-joint movements in both complete and incomplete SCI. These studies collectively reinforced the importance of engaging proprioceptive pathways for functional motor recovery.

In 2018, Wagner et al., described the first report of successful closed-loop eSCS using spatiotemporal stimulation patterns in n=3 people with incomplete SCI to enable voluntary control of previously paralyzed muscles^13^. Using this technique, they were able to enable voluntary individual joint movements such as hip, knee, and ankle flexion/extension and overground walking^13^. Most recently in 2022, Rowald et al., built on this concept of spatiotemporal stimulation patterns for n=3 participants with complete SCI. Within 1 day, participants were able to stand, walk, cycle, swim and even control trunk movements^14^. Collectively, these clinical trials have sparked significant interest within the scientific and medical communities and have encouraged the development of protocols to drive the adoption of eSCS for consistent use in patients with SCI. There are several other active clinical trials that are working to expand the use of eSCS for people living with paralysis after SCI such as the E-STAND (**Table 1**).

**Table 1.** Summary of key non-randomized clinical trials of epidural SCS for motor recovery after spinal cord injury.

| Study information |  |  | Demographics |  |  |  |  |  | SCI demographics |  |  | Rehabilitation |  | Epidural SCS |  |  |  | Safety | Outcomes |
| --- | --- | --- | --- | --- | --- | --- | --- | --- | --- | --- | --- | --- | --- | --- | --- | --- | --- | --- | --- |
| Study Name | NCT | Study region | Sample size | Age at time of study (yrs) | Age at time of injury (yrs) | Time since injury (yrs) | Sex | Upper or Lower limb | Cause of injury | AIS grade | Neurological level of injury | Type of rehab | Hours on avg | Parameters<br>Frequency (Hz) \|<br>Amplitude (V or mA) \|<br>Pulse width (µs) | Type of lead | Spinal levels targeted | Joint targeted | Adverse events | AIS Grade change: pre → post |
| Angeli et al., 2018<sup>12</sup> | NCT02339233 | USA | 4 | 25.7 (4.5) | 22.7 (4.5) | 3.1 (0.4) | M: 4<br>F: 0 | Lower limb | NR | A: 2<br>B: 2 | C5: 1<br>T1: 1<br>T4: 2 | Activity based training | 160.5 | 20-40 \| 1-6V (suprathreshold) \| 450 | Paddle | L1-S1 | hip/knee/ankle | 2; drainage, ankle | P1: A→A<br>P2: A→A<br>P3: B→C<br>P4: B→B |
| Wagner et al., 2018<sup>13</sup> | NCT02936453 | Europe | 3 | 36.7 (9.6) | 31.3 (10.7) | 5.3 (1.2) | M: 3<br>F: 0 | Lower limb | Sports: 3 | C: 2<br>D: 1 | C4: 1<br>C7: 2 | Activity based training | 160-200 | 40 \| 3-9mA (suprathreshold) \| NR | Paddle | L1-S2 | hip/knee/ankle | none | P1: C→D<br>P2: D→D<br>P3: C→C |
| Rowald et al., 2022<sup>14</sup> | NCT02936453 | Europe | 3 | 34.0 (6.2) | 29.7 (9.1) | 4.3 (4.2) | M: 3 | Lower limb | Vehicular : 3 | A: 2<br>B: 1 | T3: 1<br>T4: 1<br>T7: 1 | Activity based training | 80-100 | 20 - extensor 100 – flexor \| suprathreshold \| 300 | Paddle | L1-S2 | hip/knee/ankle | none | P1: A→A<br>P2: A→A<br>P3: B→C |

Abbreviations: AIS – American Spinal Injury Association Impairment Scale; NCT – National Clinical Trial; NR – not reported; SCS – spinal cord stimulation. Values are presented as mean (SD) or median [range], unless otherwise noted. Participants are labeled P1, P2, P3...Pn per study; bolded entries in the final column indicate a change in AIS grade over the course of the study.

### Transcutaneous SCS

#### Overview

Transcutaneous SCS emerged as a non-invasive neurostimulation alternative to eSCS. tSCS has its conceptual roots in early non-invasive percutaneous stimulation through pioneering work by Maertens de Noordhout and colleagues in the late 1980s^27^ and later by Dimitrijevic and colleagues^26^ in the late 1990s, who both established that surface stimulation over the lumbosacral spine could activate spinal sensorimotor circuits and elicit posterior root-muscle reflexes. These works fundamentally laid the groundwork for using this technology in individuals with SCI.

tSCS uses surface electrodes placed over the thoracolumbar or cervical levels (cathodes) and abdomen/iliac crests (anodes) to generate current to activate similar neural structures to eSCS^28,29^ (**Figure 1**). Oftentimes, the cathodes are placed either above and below the lesion or above and below the desired spinal levels to be targeted. Computational and pre-clinical work have supported this conceptual idea and several proof-of-concept studies have validated and supported the translation of this neurotechnology into humans^29–31^. Due to its non-invasiveness, yet relatively nuanced setup protocol, this technology has been deployed in outpatient rehabilitation clinics, where growing interest has supported pairing tSCS with task-specific training to improve voluntary motor control, standing, and upper-limb function in individuals with SCI^15,32–34^.

#### Mechanism of action

Similar to eSCS, tSCS is thought to primarily activate large-to-medium diameter Ia sensory afferent fibers within the posterior roots which then synapse onto alpha motoneurons to elicit motor responses^28^. It is assumed that for certain parameters, tSCS can in principle activate similar neural structures to enable some degree of improvement after SCI^28,31^. Nevertheless, tSCS exhibits lower selectivity for neural elements compared to eSCS, as the stimulation amplitudes of tSCS have to traverse multiple layers of skin, fat, muscle, and bone prior to reaching the spinal cord, which can compromise the ability to effectively stimulate the spinal cord^30,35^. In fact, Lieu et al., provide evidence to suggest that the effects of tSCS are largely mediated by activation of skin and muscle afferents rather than direct recruitment of dorsal root afferents^36^.

To date, tSCS has been delivered using two main waveform paradigms: low-frequency stimulation (30-50 Hz, biphasic 0.5-2ms pulses) and high-frequency carrier-modulated stimulation (5-10 kHz carrier, biphasic 0.5-1ms pulses)^15,28,32,34,37^. Conventional waveforms reliably recruit posterior root afferents but generate painful cutaneous and neuromuscular contractions limiting tolerability. Many protocols have adopted high-frequency stimulation to overcome this limitation, and have since become the dominant paradigm in the field^38–40^. However, high-frequency frequencies are known to raise the resting motor threshold, often necessitating higher stimulation amplitudes (>100mA) to recruit motor pools^38,41,42^. In fact, recent work suggests that high-frequency carrier frequencies offer no real pain benefit once the higher muscle activation thresholds are needed^35,41,42^. Moreover, Keesey et al., demonstrated that high-frequency waveforms bias recruitment toward motor efferent fibers rather than proprioceptive afferents, particularly in the cervical spinal cord, which may explain why many tSCS protocols have produced less favorable motor outcomes than anticipated^16,35^.

#### Key clinical trials

In parallel with the evolution of eSCS, a growing body of clinical trials has demonstrated that tSCS can engage similar spinal sensorimotor circuits to restore function across both the lower and upper limbs without surgical implantation. Early work by Estes et al. in 2021 evaluated the addition of thoracolumbar tSCS to locomotor training in individuals with subacute motor-incomplete SCI and demonstrated that stimulation was safe, feasible, and associated with significant improvements in walking speed and distance compared with rehabilitation alone, supporting its role as an adjunct to activity-dependent plasticity and gait recovery^34^. There has also been substantial work expanding neurostimulation to upper-extremity recovery in an attempt to better align with patients’ preferred priorities after cervical SCI. In 2021, Inanici et al. demonstrated that pairing cervical tSCS with task-specific training enabled rapid and long-term restoration of hand and arm function in individuals with chronic cervical SCI^33^. This expanded into the Up-LIFT trial, a multicenter clinical trial that evaluated tSCS in 60 participants^15^. Here, Moritz et al., showed evidence that tSCS can be used as a therapeutic neurostimulation device to restore and even sustain hand and arm function in the absence of stimulation over time, supporting its use as a practical therapy for patients in outpatient neurorehabilitation clinics.

Most recently, Bye et al. conducted the eWALK trial, the first fully powered, multicenter, triple-blind, randomized sham-controlled trial evaluating 12 weeks of tSCS combined with locomotor training in 50 individuals with chronic, motor-incomplete SCI^16^. Contrary to prior promising reports, and for the first time in a randomized trial, Bye and colleagues showed that tSCS provided no additional benefit over locomotor training alone on walking ability or any secondary outcome. These results are consistent with the findings from Keesey et al., and support the fact that high-frequency tSCS waveforms are suboptimal for meaningful recovery of motor outcomes following SCI^35^. This highlights the need for more rigorous investigation into optimal stimulation parameters, patient selection, and training protocols before tSCS can be recommended as a clinical adjunct for gait rehabilitation (**Table 2**).

**Table 2.** Summary of key non-randomized and randomized clinical trials of transcutaneous SCS for motor recovery after spinal cord injury.

| Study information |  |  | Demographics |  |  |  |  |  | SCI demographics |  |  | Rehabilitation |  | Transcutaneous SCS |  |  |  | Safety | Outcomes |
| --- | --- | --- | --- | --- | --- | --- | --- | --- | --- | --- | --- | --- | --- | --- | --- | --- | --- | --- | --- |
| Study Name | NCT | Study region | Sample size | Age at time of study (yrs) | Age at time of injury (yrs) | Time since injury (yrs) | Sex | Upper or Lower limb | Cause of injury | AIS grade | Neurological level of injury | Type of rehab | Hours on avg | Parameters: Carrier Frequency (Hz) \| Amplitude (V or mA) \| Pulse width (µs) | Lead location: Cathode \| Anode | Spinal levels targeted | Joint targeted | Adverse events | AIS Grade change: pre → post |
| Estes et al., 2021<sup>34</sup> | NCT03240601 | USA | 8 | 43.6 (18.1) | 43.4 (18.1) | 0.3 (0.1) | M | Lower limb | NR | C: 2<br>D: 6 | C1: 1<br>C2: 1<br>C3: 1<br>C4: 2<br>C5: 2<br>C7: 1 | Activity based training | 72 | NR | T11/T12 \| abdomen | NR | hip/knee/ankle | 8; mild discomfort during stimulation | NR |
| Inanici et al., 2021<sup>33</sup> | NCT03184792 | USA | 6 | 42.7 (14.2) | 38.1 (14.9) | 4.6 (3.8) | M: 4<br>F: 2 | Upper limb | NR | B: 2<br>C: 2<br>D: 2 | C3: 1<br>C5: 5 | Activity based training | 96 | 10kHz carrier \| 30 \| 40-90mA (subthreshold) \| 1000 | Above/below lesion \| iliac crest | NR | hand | 1; Mild allergic skin reaction | P1: B→B<br>P2: B→B<br>P3: C→D<br>P4: D→D<br>P5: C→C<br>P6: D→D |
| Moritz et al., 2024<sup>15</sup> | NCT04697372 | Europe, USA | 60 | 47.2 (15.5) | 41.1 (16.2) | 6.1 (7.5) | M: 50<br>F: 10 | Upper limb | Sports: 17<br>Fall: 15<br>Recreation: 14<br>Vehicular: 12<br>Other: 2 | B: 9<br>C: 28<br>D: 23 | C2: 9<br>C3: 3<br>C4: 16<br>C5: 13<br>C6: 13<br>C7: 6 | Activity based training | 25 | 10kHz carrier \| 30 \| 0-180mA (subthreshold) \| 100 | Above/below lesion \| iliac crest / clavicle.<br>Most common cathodes: C3-4/C6-7 (~50%); C4-5/C6-7 (~20%); C3-4/C7-T1 (~10%) | C3-T1 | Hand | 17 (26.6%*) events related to the device* included safety population | NR |
| Comino-Suarez et al., 2025<sup>68</sup> | NCT05210166 | Europe | 14 | 47.4 (14.5) | 46.4 (14.5) | 0.3 (0.1) | M: 14 | Lower limb | Traumatic: 7<br>Other: 8 | C: 8<br>D: 6 | C4: 6<br>C6: 1<br>T4: 1<br>T6: 1<br>T9: 1<br>T10: 4 | Activity based training | 20 | No carrier \| 20 \| subthreshold \| 1000 | T11-T12 \| umbilicus | L1-S1 | hip/knee/ankle | 14 (100%): Skin irritation ; 12 (85.6%) : discomfort during stimulation | NR |
| Bye et al., 2026<sup>16</sup> | ACTRN12620001241921 | Australia, Europe, USA | 25 | 46 [IQR: 37-56] | NR | > 1 year | M: 20<br>F: 5 | Lower limb | NR | A: 4<br>B: 0<br>C: 16<br>D: 5 | C7-T5: 11<br>T6-T11: 14 | Activity based training | 18 | 10kHz carrier \| 20 \| subthreshold \| 1000 | L1-2 \| abdomen | L1-S1 | hip/knee/ankle | 56 events; 22 (39.2%) skin irritation ; 6 (10.7%) increase spasticity; 6 | NR |

### Therapeutic goals

Although eSCS and tSCS engage similar spinal circuits, they are designed to serve fundamentally distinct therapeutic goals.

Briefly, eSCS is designed as a single, permanently implanted neurostimulation system intended as an assistive device that elicits immediate improvements in motor function as soon as stimulation is turned on. Typically, following the procedure, eSCS is used in conjunction with intensive rehabilitation to maximize the potential of recovery. After rehabilitation, stimulation is designed to be continued in the at-home setting with parameters programmed in the initial rehabilitation setting. Since the electrodes are secured into place and with constant parameters, functional gains can be reproduced and sustained as soon as stimulation is turned on. Thus, the therapeutic benefit of eSCS is designed to be with continuous use over time.

In contrast, tSCS is primarily designed as a therapeutic neurostimulation device that is used in the outpatient clinical rehabilitative setting with its main therapeutic benefit emerging after weeks of rehabilitation, rather than as an immediate assistive effect. Furthermore, tSCS is best used in the rehabilitation setting with experienced providers who are able to place electrodes precisely and tune the stimulation parameters, thereby limiting its feasibility for independent at-home use.

## RESULTS

To address the fundamental clinical questions of when clinicians should choose eSCS or tSCS, and who are the responders to either technique, we conducted a systematic literature search and pooled together individual participant data to explore these differences in depth and define indications based on their intended use and the durability of their effects.

### Study selection

The published literature was systematically searched in PubMed, EMBASE, and Web of Science from database inception through April 2026, using the keywords “epidural spinal cord stimulation,” “transcutaneous spinal cord stimulation,” “spinal cord injury,” “motor recovery,” and “rehabilitation,” combined with related MeSH terms. Complete search strategies are provided in the **Supplementary Table 1**.

Studies included in this Review met the following inclusion criteria: (i) reported *de novo* participant-level data from human participants with SCI receiving eSCS or tSCS; (ii) reported quantitative upper or lower limb motor outcomes, including EMG activity, single joint movement, walking, or standing; and (iii) were published in English. Studies were excluded if they: (i) included in-vitro or animal data; (ii) contained overlapping participants from previously included studies; iii) were systematic reviews, meta-analyses, editorials, or letters lacking primary data.

Of the 1547 studies identified across databases, 407 duplicates were removed, leaving 1140 studies for title and abstract screening. Of these, 97 underwent full-text screening, of which 52 ultimately fit inclusion criteria. Of these 52 included studies, 44 (26 eSCS and 18 tSCS) reported *de novo* participant-level data and were incorporated into the pooled cohort analysis (**Supplementary Tables 2-5**).

### Pooled cohort

From this corpus, the 44 included studies yielded data from 278 unique participants, of whom 94 received eSCS and 184 received tSCS. Participants in eSCS studies were significantly younger both at the time of the study enrollment (32.1±10.3 vs. 42.1±15.9, p<0.0001) and at the time of the injury (27.4±9.9 vs. 35.8±16.0, p<0.0001). Furthermore, both cohorts had similar distributions of sex, with a male predominance (78.7% eSCS vs. 83.7% tSCS).

Subgroup analyses showed differences between cohorts treated with eSCS and tSCS. Studies of tSCS predominantly enrolled participants with motor incomplete injuries (AIS C-D, eSCS: 8.6% vs. tSCS: 78.8%, p<0.001), whereas eSCS studies predominately enrolled participants with motor complete injuries (AIS A-B, eSCS: 91.4% vs. tSCS: 21.2%).

In parallel, injury level differed substantially between modalities. Participants receiving tSCS were far more likely to have cervical injuries compared with those in eSCS studies (eSCS: 36.2% vs. tSCS: 78.6%, p<0.001). Most studies targeted participants in the chronic phase of recovery (eSCS: 97.9% vs. tSCS: 76.7%), although populations treated in the subacute phase were more commonly enrolled in tSCS studies (eSCS: 2.1% vs. tSCS: 23.3%). Lastly, the therapeutic targets of stimulation diverged markedly: tSCS studies primarily focused on upper-limb impairments (52.7%), whereas eSCS studies almost exclusively targeted lower-limb function (97.9%), irrespective of the level of injury (**Figure 2, Supplementary Table 6**).

**Fig. 2.**
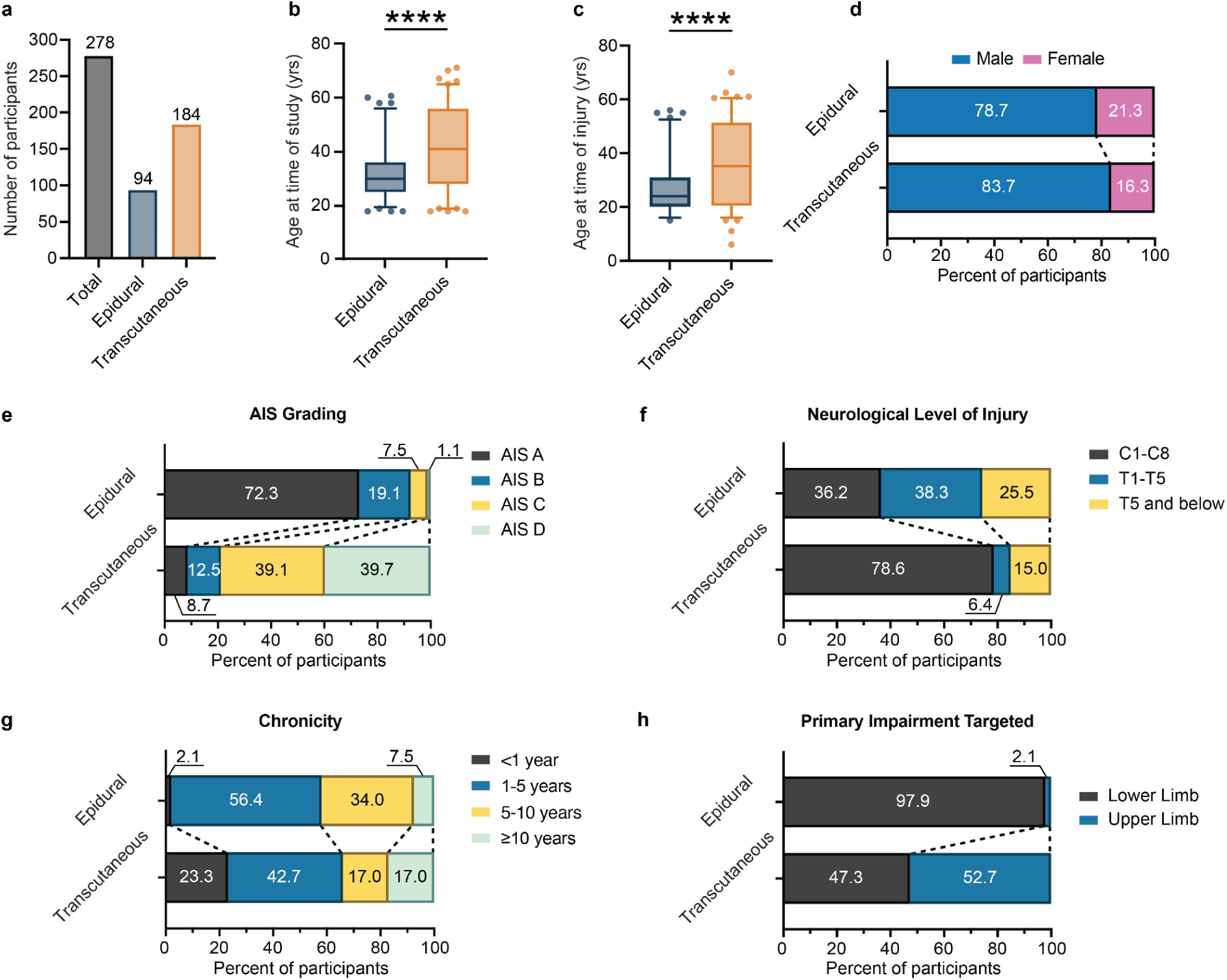
Demographics of the participants among included studies. **a.** Total number of participants included, stratified by epidural and transcutaneous SCS. **b.** Age at the time of study. **c.** Age at the time of injury. **d-h.** Bar plots of various variables stratified by epidural and transcutaneous SCS across various distributions including **d.** sex; **e.** AIS status at study; **f.** neurological level of injury; **g.** chronicity; and **h.** primary impairment targeted.

### SCS parameters

The parameters used for participants are one of the most important aspects for effectively translating this technology into clinical settings. However, tuning parameters is a tedious process and determining which specific parameters to use also remains elusive. Furthermore, traditional parameter tuning has been mainly based on prior anecdotal experience, and the literature currently lacks standardized in-human guidelines to encourage systematic programming of these participants. To help address these concerns, we pooled together the parameters used across all studies in this Review (**Figure 3, Supplementary Table 6**).

**Fig. 3.**
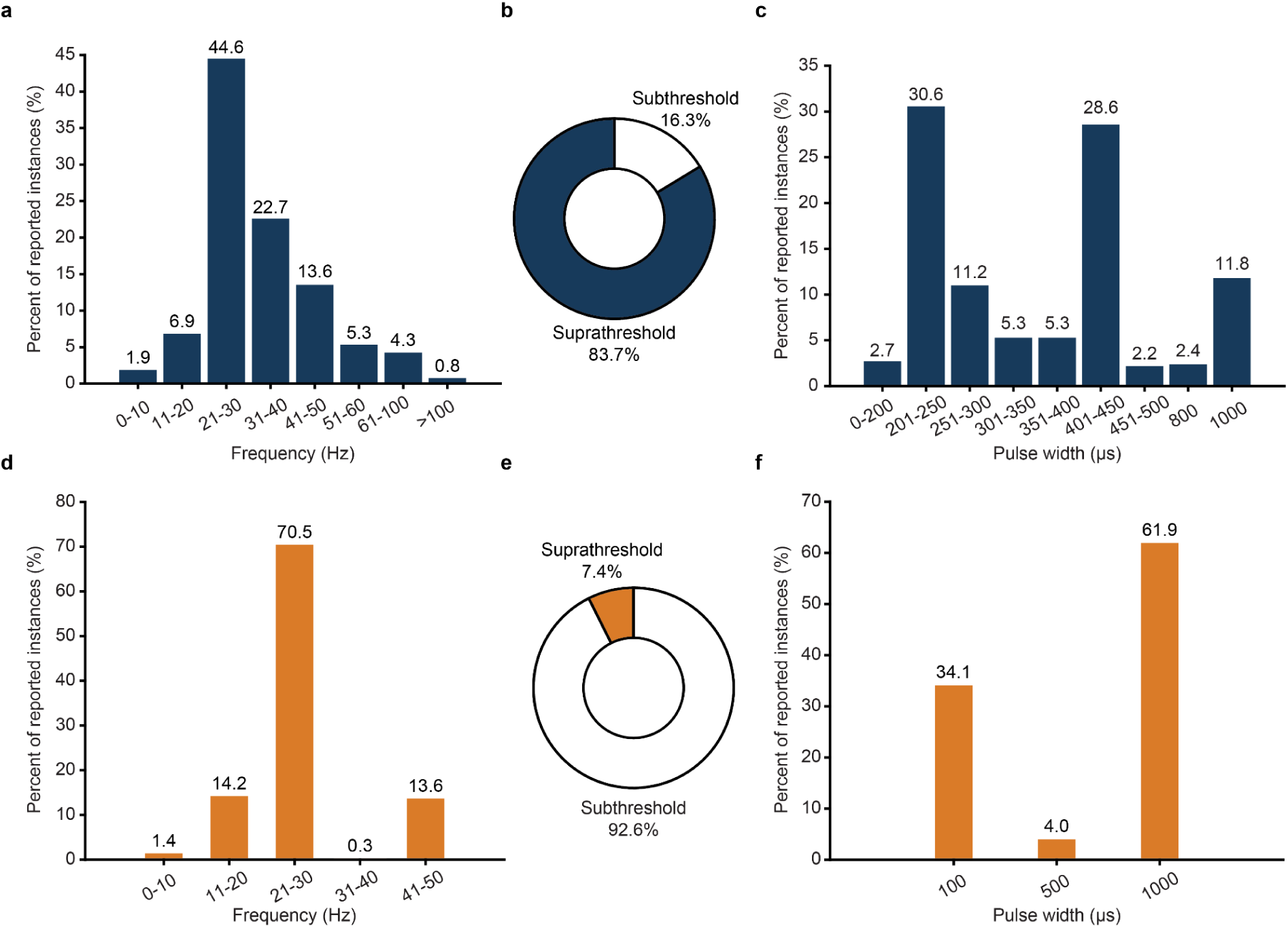
SCS parameters a-c. Epidural SCS parameters. **a.** Histogram of the frequencies (Hz) used in studies with epidural SCS. Frequencies are binned according to distinct ranges, and the percentage of all reported instances are plotted. **b.** Distribution of supra- and submotor threshold. **c.** Histogram of the pulse widths (µs) used in studies with epidural SCS. Pulse widths are binned according to distinct ranges, and the percentage of all reported instances are plotted**. d-f.** Transcutaneous SCS parameters. **d.** Histogram of the frequencies (Hz) used in studies with transcutaneous SCS. Frequencies are binned according to distinct ranges, and the percentage of all reported instances are plotted. **e.** Distribution of supra- and submotor threshold. **f.** Histogram of the pulse widths (µs) used in studies with transcutaneous SCS. Pulse widths are binned according to distinct ranges, and the percentage of all reported instances are plotted.

#### Parameters: epidural and transcutaneous SCS

The majority of participants in the eSCS cohorts were stimulated within a range of 21-50Hz (80.9%). Additionally, the majority of amplitudes stimulated was in the supramotor threshold range (83.7%). However, there was a significant distribution on the pulse width, with several frequency peaks appearing at 200µs, 450µs, and 1000µs (30.6%, 28.6%, 11.8%, respectively) (**Figure 3a-c, Supplementary Table 6**).

Among the included studies, tSCS was most commonly delivered between 21-50Hz (84.4%), following a similar rate to eSCS. However, contrary to the eSCS cohorts, the majority of stimulation amplitudes were predominately applied at submotor threshold (92.6%), typically exceeding sensory threshold with a range of 16-180mA. Pulse widths were discretely clustered at 100, 500, and 1000µs, representing 34.1%, 4.0%, 61.9% of cases respectively (**Figure 3d-f, Supplementary Table 6).**

#### Tailoring stimulation to upper vs. lower limb in transcutaneous SCS

Furthermore, when stratifying between cervical and lumbar applications, stimulation parameters differed. For participants receiving cervical tSCS, frequencies between 21-30 Hz were used in nearly all instances (97.9%), with 41-50 Hz reported in only 2.1% of cases. Pulse widths used were primarily 100µs (61.9%), followed by 1000µs (30.9%) and 500µs (7.2%). In contrast, lumbar tSCS more frequently targeted 21-30Hz (37.3%), followed by 11-20Hz (31.6%), and 41-50Hz (26.6%), with 1000µs representing the only pulse width reported. Submotor threshold amplitudes were applied with similar rates in both regions (92.8% in cervical vs. 93.7% in lumbar stimulation). Additional details are provided in **Supplementary Table 6**.

### Responder analysis

Despite increasing evidence supporting the role of SCS for facilitating motor rehabilitation outcomes for people living with SCI, there remains inconsistent evidence to suggest the determinant of responsiveness following either eSCS or tSCS. To address these limitations, we aimed to integrate the pooled data to better define characteristics of responsiveness following both eSCS and tSCS. Here, we focused on comparing two important biomarkers for recovery: voluntary EMG activity and voluntary control of single joint movements. These biomarkers were consistently reported across both eSCS and tSCS studies, allowing for the most direct comparisons. Furthermore, to evaluate the extent of therapeutic benefit of eSCS and tSCS, we were interested in two key metrics: response to therapy, and the time taken to observe the first improvements in each, which we evaluated through a cumulative incidence survival analysis.

#### Determinants of response

When evaluating the outcomes to recovering voluntary EMG activity, eSCS showed remarkably consistent results. All participants responded to eSCS despite the extent of their injury (AIS A-D), while tSCS appeared to only consistently benefit individuals with incomplete motor injuries (AIS C-D) (**Figure 4a**). To further evaluate the time to response of the therapy, we found that eSCS demonstrated superiority compared to tSCS (**Figure 4b**, p<0.001), with eSCS showing >50% of response within 1 week and 90% of response within 14 weeks. tSCS participants took a longer time to recover voluntary EMG activity, with a 50% response rate at 18 weeks. These results were also similar when evaluating functional movements, such as a regained or improved single joint movement (**Figure 4c-d**, p<0.0001).

**Fig. 4.**
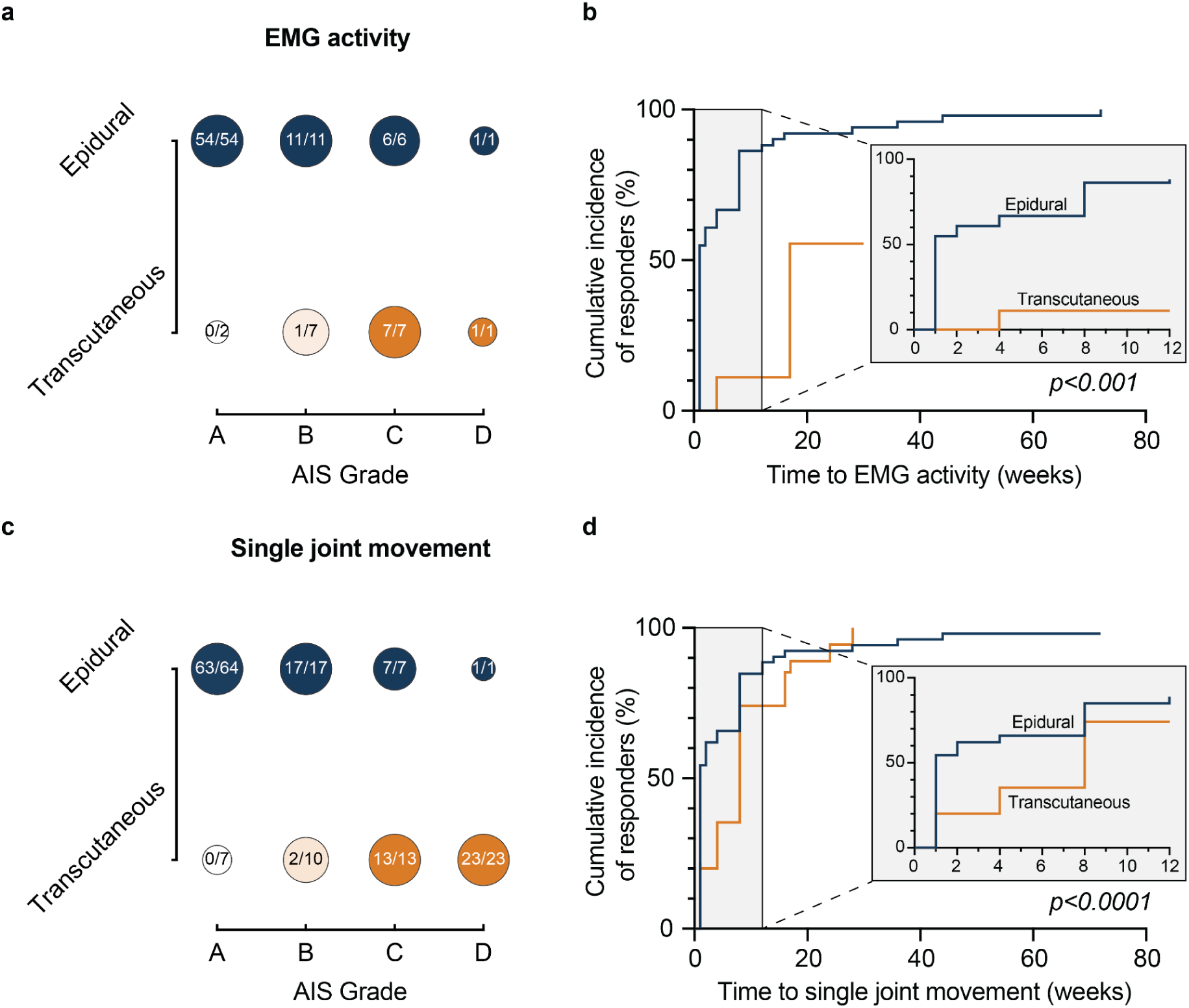
Determinants of response to epidural and transcutaneous SCS following volitional activity. **a.** Dot plot representing the number of responders with recovery of EMG activity during volitional tasks, stratified across AIS status. The size of the dot represents the proportional sample size in each AIS subcategory, and the shade of the color represents the frequency of the occurrence. **b.** Cumulative time to recovery of volitional EMG activity showing that the majority of eSCS cohorts achieve recovery of EMG activity faster compared to the tSCS cohorts (p<0.001). **c.** Dot plot representing the number of responders with recovery of single joint activity, stratified across AIS status. The size of the dot represents the proportional sample size in each AIS subcategory, and the shade of the color represents the frequency of the occurrence. **d.** Cumulative time to recovery of volitional single joint movement, showing that the majority of eSCS cohorts achieve volitional single joint movements at a faster rate compared to the tSCS cohorts (p<0.0001).

Furthermore, we stratified cohorts based on AIS status. Here we observed that eSCS is superior for individuals with both AIS A and AIS B-D (**Supplementary Figure 1**). Importantly, when controlling for the amount of stimulation dose, eSCS also demonstrated faster recovery times (**Supplementary Figure 2**). Furthermore, eSCS can elicit improvements in motor function despite the degree of injury, while tSCS requires spared sensory function in order to regain functional single joint movements (**Supplementary Figure 1b,d**).

### Safety

Overall, the safety profile of both eSCS and tSCS remained strong, with both modalities reporting no serious adverse events associated with stimulation. Across the 94 participants implanted with eSCS, adverse events were rare (total adverse event rate: 7.4%), including two urinary tract infections (2.1%), two cases of lead migration (2.1%), one case of hip fracture (1.1%), one instance of IPG site drainage (1.1%), and one case of ankle edema (1.1%). Comparatively, among the tSCS cohort, there were a total of 111 (60.3%) reported mild or transient adverse events. These included mild skin irritation or blistering (28.8%), intense tingling during stimulation (11.9%), worsened spasticity (4.3%), mild urinary incontinence (2.2%), autonomic dysreflexia (1.1%), nausea (0.5%), and other unspecified minor events (11.5%).

## DISCUSSION

Over the past two decades, eSCS and tSCS have emerged as promising neurostimulation modalities for recovering voluntary motor function after SCI. Despite growing evidence to support both approaches, clinicians and researchers alike have lacked a clear framework to distinguish these two approaches. Four fundamental questions remain unaddressed: when should eSCS or tSCS be chosen, how do the therapeutic goals of each modality differ, what stimulation configurations maximize benefit, and who are the ideal responders for each technique? In this Review, we addressed each of these questions to provide a rational, evidence-based framework to guide the clinical adoption of eSCS and tSCS.

### Who are the populations targeted for eSCS and tSCS?

Within the pooled cohort, we found that participants in eSCS studies were significantly younger both at the time of the study enrollment and at the time of the injury. This age difference may likely reflect selection biases associated with surgical candidacy and willingness to pursue invasive interventions. Furthermore, both eSCS (78.7%) and tSCS (83.7%) cohorts had a higher proportion of male participants, which reflects the current epidemiology of SCI within the USA, in which approximately 78% of affected individuals are male^2^. Additionally, tSCS predominantly enrolled participants with motor incomplete injuries (AIS C-D, 78.8%), while eSCS predominately enrolled those with motor complete injuries (AIS A-B, 91.4%). This likely reflects how current rehabilitation paradigms are structured, in which individuals with motor incomplete injuries, who retain greater spontaneous recovery potential, are preferentially directed for noninvasive, rehabilitation-based interventions such as tSCS, whereas eSCS has been reserved for those with more severe injuries and limited alternative therapeutic options^43^.

However, perhaps the most striking finding was the marked difference in therapeutic purpose between tSCS and eSCS for individuals with cervical SCI. While 71% of cervical SCI participants who received tSCS were targeted for upper limb recovery, only 5.9% of cervical SCI participants in eSCS studies were intentionally targeted for the upper limb. This discrepancy is notable given that patient-reported priorities in cervical SCI rehabilitation consistently demonstrate that restoration of upper limb function is valued above all other domains of daily living, including sexual function, trunk stability, bowel and bladder control, ambulation, sensation, and pain^43^.

This difference is likely due to historical emphasis of eSCS on lower limb outcomes that reflected early conceptual frameworks centered on spinal central pattern generators (CPGs) which have locomotor behavioral patterns and are believed to operate largely independently of supraspinal input^26^. However, emerging evidence suggests that despite the lack of CPGs within cervical spinal circuits, these are still amenable to effective modulation. Notably, in 2016, Lu et al. demonstrated for the first time that cervical eSCS could meaningfully improve gross motor function in two individuals with AIS B injuries^44^. Furthermore, in the largest clinical trial to date encompassing these technologies, Moritz et al., described findings supporting the use of tSCS for the recovery of upper limb function in rehabilitation clinics^15^. At the moment, tSCS studies currently align more with patients’ priorities compared to those involving eSCS.

### What are the differences in the therapeutic goals of eSCS and tSCS?

There are distinct usability considerations for both eSCS and tSCS which influence how each approach is deployed clinically. Once implanted and programmed, eSCS can be used independently by patients in home environments, allowing flexible selection of stimulation programs, waveforms, and contacts to match functional goals during daily activities^13,14^. This supports long-term, continuous integration of stimulation into rehabilitation and everyday living. In contrast, tSCS requires careful electrode placement, parameter adjustment, and often produces discomfort during stimulation, making routine independent use challenging. Additionally, the setup is tedious and typically requires trained personnel, limiting its practical use to supervised clinical or laboratory rehabilitation sessions. Furthermore, we found that eSCS has an assistive effect that is achieved as soon as stimulation is turned on and is sustained over time (**Figure 4**). In contrast, we found that tSCS demonstrates a therapeutic effect that emerges after weeks of concurrent rehabilitation, rather than as an immediate assistive effect (**Figure 4**). As a result, current clinical implementation positions eSCS as an immediate and long-term neurostimulation strategy, whereas tSCS is better suited as an interval therapeutic rehabilitation adjunct.

### What are the optimal stimulation configurations for eSCS and tSCS?

To enable rapid and reliable clinical adoption of eSCS and tSCS, stimulation configurations for both eSCS and tSCS must be close to standardized. While prior work has established stimulation protocols in animal models, comparable in-human studies remain limited^45^. As a result, parameters have historically been guided using anecdotal experience rather than systematic evidence. To address this gap, we pooled the stimulation parameters reported across all studies included in this Review (**Figure 3, Supplementary Table 6**).

#### Frequency: Why 20-50Hz?

Notably, we found that the majority of stimulation of participants in both eSCS (80.9%) and tSCS (84.4%) cohorts were stimulated within the 21-50Hz range, which raises questions of why this frequency band has been consistently chosen for both modalities. Selection of stimulation frequency in eSCS and tSCS studies for motor recovery is grounded in early physiological studies demonstrating frequency-dependent recruitment of spinal locomotor networks^26,46,47^. Unlike traditional SCS systems designed for pain control, which often employs very high-frequencies (>1kHz) to suppress proprioceptive fibers, SCS designed for motor recovery leverages lower frequencies to engage intrinsic spinal circuitry. In 1973, Cook et al., first observed that low frequency stimulation facilitated motor function in a patient with multiple sclerosis^19^. Thereafter, in 1998, Dimitrijevic and colleagues reported that lumbar stimulation at 25-60Hz generated rhythmic flexion/extension patterns in six paraplegic individuals and proposed this finding as functional evidence of CPGs in humans^26^. This led to a number of studies exploring low frequency stimulation of the lumbosacral segments to facilitate voluntary motor control. Harkema et al., described that stimulation at 15Hz preferentially facilitated standing, whereas higher frequencies in the 30-40Hz range promoted stepping^11^. This was further replicated by a number of studies suggesting slightly nuanced stimulation differences to facilitate specific functional improvements such as walking, standing, and cycling^12–14,48–50^. This has also been the experience in tSCS studies to improve grasp/grip function^15,32,33^. Further work by Wagner et al., explored the role of leveraging the fact that flexor and extensor motor pools respond preferentially to distinct frequencies, with lower frequencies (20Hz) activating extensors while higher frequencies (100Hz) preferentially recruiting flexor motor pools^13,14,51^. More recent approaches by Romeni et al., describe similar philosophies of differential frequencies by combining low-frequency (40Hz) with high-frequency stimulation (1.2kHz) to reduce pathological co-contraction which has shown to replicate physiological motor behavior^48^. Despite this progress, there is still limited robust evidence that explains why certain frequency ranges are more efficacious from a mechanistic standpoint. Thus, this is an area of strong scientific interest that future work should aim to address.

#### Stimulation amplitudes

Interestingly, the majority of stimulation amplitudes for eSCS studies were generally suprathreshold **(Figure 3b)**. In contrast, tSCS was predominantly delivered at subthreshold amplitudes **(Figure 3e)**. It is important to note that subthreshold stimulation at rest can generate suprathreshold responses when combined with volitional motor drive^24^. This may be explained by the fact that the majority of participants who were implanted and evaluated by eSCS were motor complete injuries (ASIA A-B, 92.5%) where suprathreshold stimulation is necessary to facilitate voluntary motor movement in previously dormant neural circuits downstream of the injury.

### Who are the ideal responders to eSCS and tSCS?

In a 2022 review, Seáñez and Capogrosso proposed that intact sensory function (ie. AIS B-D) may predict response in eSCS across various domains such as voluntary EMG activity, single joint movements, as well as functional improvements such as treadmill and overground walking and standing^52^. However, that study was limited to just 8 participants across 3 studies which limited its generalizability. Here, we extend their work by incorporating a substantially larger population of participants from the most current literature, while directly comparing eSCS and tSCS to identify the ideal responder profile for both modalities.

Our findings, albeit strong, were not surprising. eSCS produces an assistive effect that emerges considerably faster than tSCS (1 week vs. 18 weeks for restoring voluntary muscle activity). This effect is sustained over time with continued stimulation. Perhaps the strongest results are supporting the use of eSCS for all participants despite the extent of their injury (AIS A-D, **Figure 4a**). Previously, options for individuals with motor complete injuries were limited; however, these results show promising evidence that eSCS has a meaningful role in effective rehabilitation for even the most severe injury subpopulations. In contrast, tSCS appears most beneficial for individuals in motor incomplete cohorts (AIS C-D), with improvements emerging only after stimulation periods of up to 18 weeks (**Figure 4b,c**).

Previously, it was hypothesized that the duration of stimulation has an effect on the resulting motor improvements, similar to a rehabilitation dose that is given in clinics. It is clear with this data that even when correcting for the total hours of stimulation, eSCS enables response substantially more rapidly compared to tSCS (**Supplementary Figure 2**). This supports eSCS as a neurostimulator that produces an assistive effect that emerges immediately as soon as stimulation is turned on. It is important to note, that if stimulation is withdrawn for eSCS, participants will indeed worsen. Therefore, it is necessary to sustain stimulation to maintain its therapeutic effects. Meanwhile, tSCS is best suited as a therapeutic neurostimulation device that offers the best outcomes when paired with rehabilitation.

### What other functional gains were observed following eSCS and tSCS?

Although our analysis focused on volitional recovery of EMG activity and single-joint movements, the most clinically meaningful outcomes are improvements in functional capacity measured using validated clinical scales. In this analysis, however, direct head-to-head comparisons were limited by heterogeneous data.

In eSCS cohorts, AIS motor scores increased as much as +23 points for participants undergoing upper limb eSCS^44^, and +16 for participants undergoing lower limb eSCS^13^. Notably, across eSCS studies that reported changes in AIS grading and neurological level of injury status, 13 participants experienced a clinically meaningful improvement of at least one AIS grade (ie. A→D or C→D), and 9 participants demonstrated an improvement in the neurological level of injury of at least one spinal segment. Interestingly, the majority of participants who improved in at least one AIS grade were in the Kandhari study, where they enrolled participants in the subacute period^53^. In the absence of a control group, it is difficult to determine the extent to which these gains can be attributed to stimulation alone versus spontaneous recovery. Nonetheless, these findings provide preliminary evidence that pairing eSCS with rehabilitation in the subacute period may potentiate substantial improvements in overall function, including rare transitions from AIS A→D.

In the largest tSCS clinical trial systematically reporting these outcomes, Moritz et al. demonstrated gains across strength, functional, and sensory domains in individuals with chronic cervical SCI^15^. While improvements in strength during the tSCS period were observed for GRASSP Strength (+2.8) and Prehension Ability (+1.6), these effects were also seen during the rehabilitation period, making the exercise the participants engaged in a confounding factor. In contrast, sensory recovery appeared to be the largest meaningful effect of tSCS. ISNCSCI total sensory score did not improve during rehabilitation alone but increased by +9.6 points during the tSCS period, suggesting that sensory recovery was driven by stimulation rather than exercise. Furthermore, of all the tSCS studies that reported changes in AIS grading and level of neurological injury, 6 participants improved by at least one AIS grade^33,39^ and one improved by at least one spinal level^32^.

### Future directions for eSCS and tSCS

Although the effects following eSCS or tSCS use after SCI show remarkable improvements in motor function during and after stimulation, these technologies were mainly focused on assisting patients in the chronic phase of their recovery. This was the primary focus early on to establish SCS as a feasible therapy. However, it is well known that the effects of spontaneous recovery occur within the subacute phases of recovery^6,54^. The subacute phase may represent a critical time window where the maximum effects of recovery are possible^55^. In fact, among the studies included in our analysis, only 2.1% and 23.3% of participants for eSCS and tSCS, respectively, intervened during the subacute phase of their recovery. These studies showed similar improvements with concurrent rehabilitation, which is promising^34,53,56^. As a result, logical next steps would be to intervene more consistently with eSCS and tSCS in the subacute period in order to maximize the potential for recovery after SCI.

Additionally, the versatility of SCS may expand beyond initial indications of restoring motor function and into other important sequelae of SCI such as autonomic functions, including hemodynamic instability, sexual function, and bowel and bladder function^43^. This has been a recent focus for research groups to design therapeutics that can restore some of these functions and diminish the maladaptive effects of SCI, including orthostatic hypotension^57^, autonomic dysreflexia^58^, bowel and bladder incontinence^59,60^, and even restore sexual function^61,62^. Importantly, this has led to a pivotal trial (EMPOWER-BP) to explore these effects further in a large multi-institutional cohort^57^.

Lastly, future clinical trials must focus on addressing patient-specific priorities. For example, restoration of upper limb function remains the top goal for individuals with cervical SCI, whereas restoration of autonomic function remains the top priority for those with thoracic or lower injuries^43^. Tailoring stimulation paradigms to injury level and patient-reported functional goals will certainly be essential for ensuring strong clinical translation, maximizing long-term functional independence after SCI, and ultimately achieving buy-in from patients, clinicians, and scientists alike. Beyond SCI, SCS has the potential to impact many other neurological disorders that cause motor deficits across the lifespan, from pediatric to adult populations. These include neurovascular injuries such as stroke^63–65^, and neurodegenerative diseases such as spinal muscular atrophy^66^, Parkinson’s disease^67^, amyotrophic lateral sclerosis and cerebral palsy.

## CONCLUSION

SCS has emerged as a promising neurostimulation technology capable of restoring voluntary movement across the full spectrum of SCI severity. Over the past two decades, both eSCS and tSCS have seen increasing adoption in human cohorts. In this Review, we summarized the unique characteristics of participants enrolled in these studies to date and identified potential determinants of response for each technique. eSCS has emerged and is likely to remain as an assistive technology that can improve function immediately upon stimulation with gains persisting over time with continuous stimulation. With eSCS, participants recover volitional muscle activity and single joint movements within 1 week of stimulation and respond despite the extent of their injury (AIS A-D). In contrast, tSCS is a technology that is currently designed for long-term therapeutic intervention with concurrent rehabilitation. tSCS demonstrates cumulative gains that emerge over periods of up to 18 weeks, particularly benefitting those with motor incomplete injuries (AIS C-D).

## Data Availability

All data produced in the present work are contained in the manuscript

## Notes

### Competing Interest Statement

The authors have declared no competing interest.

### Author Declarations

The study used only openly available human data that were obtained from studies from our systematic literature search. All data were anonymized.

