## Supplementary Materials for "Epidural Versus Transcutaneous Spinal Cord Stimulation For Motor Recovery After Spinal Cord Injury: A Comparative Analysis"

### Supplementary Table 1. Search strategy

| Database | Search strategy |
|---|---|
| **PubMed** | ("Spinal Cord Stimulation"[Mesh] OR "Epidural Stimulation" OR "Spinal Cord Neuromodulation" OR "SCS" OR "Transcutaneous Stimulation") AND ("Spinal Cord Injuries"[Mesh] OR "SCI" OR "Paralysis"[Mesh] OR "Hemiparesis" OR "Paraplegia"[Mesh] OR "Tetraplegia") AND ("Motor Recovery" OR "Motor Function" OR "Motor Improvement" OR "Functional Recovery" OR "Rehabilitation"[Mesh]) NOT (Review[pt] OR "Systematic Review"[pt] OR "Meta-Analysis"[pt]) |
| **EMBASE** | ('spinal cord stimulation'/exp OR 'epidural stimulation' OR 'spinal cord neuromodulation' OR 'SCS' OR 'transcutaneous stimulation' OR 'transcutaneous spinal stimulation' OR 'tSCS') AND ('spinal cord injury'/exp OR 'SCI' OR 'paralysis'/exp OR 'hemiparesis' OR 'paraplegia'/exp OR 'tetraplegia') AND ('motor recovery' OR 'motor function' OR 'motor improvement' OR 'functional recovery' OR 'rehabilitation'/exp) NOT ('systematic review'/it OR 'meta analysis'/it OR 'review'/it) |
| **Web of Science** | TS=("Spinal Cord Stimulation" OR "Epidural Stimulation" OR "Spinal Cord Neuromodulation" OR "SCS" OR "Transcutaneous Stimulation" OR "Transcutaneous Spinal Stimulation" OR "tSCS") AND TS=("Spinal Cord Injury" OR "SCI" OR "Paralysis" OR "Hemiparesis" OR "Paraplegia" OR "Tetraplegia") AND TS=("Motor Recovery" OR "Motor Function" OR "Motor Improvement" OR "Functional Recovery" OR "Rehabilitation") |

### Supplementary Table 2. Summary of non-randomized clinical trials of epidural SCS for motor recovery after spinal cord injury

| Study information | | | Demographics | | | | | | SCI demographics | | | Rehabilitation | | Epidural SCS | | | | Safety | Notes |
|---|---|---|---|---|---|---|---|---|---|---|---|---|---|---|---|---|---|---|---|
| **Study Name** | **NCT** | **Study region** | **Sample size** | **Age at time of study (yrs)** | **Age at time of injury (yrs)** | **Time since injury (yrs)** | **Sex** | **Upper or Lower limb** | **Cause of injury** | **AIS grade** | **Neurological level of injury** | **Type of rehab** | **Hours on avg** | **Parameters Frequency (Hz) \| Amplitude (V or mA) \| Pulse width (µs)** | **Type of lead** | **Spinal levels targeted** | **Joint targeted** | **Adverse events** | |
| Angeli et al., 2018[12] | NCT02339233 | USA | 4 | 25.7 (4.5) | 22.7 (4.5) | 3.1 (0.4) | M: 4 F: 0 | Lower limb | NR | A: 2 B: 2 | C5: 1 T1: 1 T4: 2 | Activity based training | 160.5 | 20-40 \| 1-6V (suprathreshold) \| 450 | Paddle | L1-S1 | hip/knee/ankle | 2; drainage, ankle | |
| Wagner et al., 2018[13] | NCT02936453 | Europe | 3 | 36.7 (9.6) | 31.3 (10.7) | 5.3 (1.2) | M: 3 F: 0 | Lower limb | Sports: 3 | C: 2 D: 1 | C4: 1 C7: 2 | Activity based training | 160-200 | 40 \| 3-9mA (suprathreshold) \| NR | Paddle | L1-S2 | hip/knee/ankle | none | |
| Darrow et al., 2019[61] | NCT03026816 | USA | 2 | 50.0 (2.8) | 42.5 (0.7) | 7.5 (3.5) | M: 0 F: 2 | Lower limb | Fall: 1 Vehicular: 1 | A: 2 | T4: 1 T8: 1 | NR | NR | 24-50 \| 10-16mA (suprathreshold) \| 420 | Paddle | L1-S2 | hip/knee/ankle | none | |
| Calvert et al., 2019[*69] | NCT02592668 | USA | 1* | 37 | 31 | 6 | M | Lower limb | Fall | A | T3 | Activity based training | 72 | 40 \| <7V (suprathreshold) \| 210 | Paddle | L1-S1 | hip/knee/ankle | none | Continuation of Gill et al., 2018 |

| | | | | | | | | | | | | | | | | | | | |
|---|---|---|---|---|---|---|---|---|---|---|---|---|---|---|---|---|---|---|---|
| Pena Pino et al., 2020*[70] | NCT03026816 | USA | 5* | 40 [30-60] | NA | 7.5 (5.5) | M: 4 F: 1 | Lower limb | Sports: 2 Vehicular: 2 Fall: 1 | A: 4 B: 1 | T4: 2 T5: 2 T8: 1 | Activity based training | 125 | 16-400 \| 2-15mA (suprathreshold) \| 200-500 | Paddle | L2-S2 | hip/knee/ankle | none | Continuation of Gill et al., 2018 and Darrow et al., 2019 |
| Beck et al., 2021*[71] | NCT02592668 | USA | * | - | - | - | - | - | - | - | - | - | - | - | - | - | - | UTI | Same participant as Calvert et al., 2019 |
| Gill et al., 2021*[72] | NCT02592668 | USA | * | - | - | - | - | - | - | - | - | - | - | - | - | - | - | UTI | Same participant as Calvert et al., 2019 |
| Rowald et al., 2022[14] | NCT02936453 | Europe | 3 | 34.0 (6.2) | 29.7 (9.1) | 4.3 (4.2) | M: 3 | Lower limb | Vehicular: 3 | A: 2 B: 1 | T3: 1 T4: 1 T7: 1 | Activity based training | 80-100 | 20 - extensor 100 – flexor \| suprathreshold \| 300 | Paddle | L1-S2 | hip/knee/ankle | none | |
| Gorgey et al., 2023 | NCT04782947 | USA | 2 | NR | NR | 2 (2.1) | M: 2 | Lower limb | NR | A: 1 B: 1 | C8: 1 T11: 1 | Activity based training | 144 | 30 \| suprathreshold \| 450 | Perc | L1-S1 | hip/knee/ankle | None | |
| Singh et al., 2023[73] | NCT03026816 | USA | 2 | 25 [20-30] | NR | 2.0 (4.9) | M: 2 | Lower limb | NR | A: 2 | T4: 1 T6: 1 | NR | NR | 28-44 \| 2.9-16 (suprathreshold) \| 400-500 | Paddle | L1-S1 | hip/knee/ankle | None | Continuation of Darrow et al., 2019, Pena Pino et al., 2020. |
| Wan et al., 2024[74] | NCT05644171 | Asia | 2 | 36.5 (14.8) | 31.0 (14.1) | 5.5 (0.7) | M: 2 | Lower limb | Sports: 1 Vehicular: 1 | A: 2 | T1: 1 T4: 1 | Activity based training | 140-210 | 50 \| 5-20mA (suprathreshold) \| 100-400 | Paddle | L1-S1 | hip/knee/ankle | none | |
| Romeni et al., 2025[48] | NCT05926843 | Europe | 2 | 43.5 (16.3) | 41.2 (15.3) | 2.3 (0.9) | M: 1 F: 1 | Lower limb | Sports: 1 Vehicular: 1 | C: 2 | T3: 1 T5-7: 1 | NR | NR | Low, High Freq Stimulation: 40, 1200 \| subthreshold \| 300, 20µs | Paddle (32 channel) | L1-S1 | hip/knee/ankle | none | |
| Rybka et al., 2025[51] | NCT05690074 | Europe | 3 | 29.8 (4.1) | 26.5 (3.4) | 3.2 (0.9) | M: 3 | Lower limb | Vehicular: 2 Sports: 1 | A: 3 | T2: 1 T4: 2 | Activity based training | >100 | 20 - extensor 100 – flexor \| 2-15mA (suprathreshold) \| 210-400 | Paddle (32 channel) | L1-S1 | hip/knee/ankle | 2; ileus. IPG migration within subcutaneous tissue. Developed pressure ulcer. Required re-exploration of IPG | |
| Wee et al., 2025*[75] | NCT05644171 | Asia | 1* | 38 | 27 | 11 | F | Lower limb | Fall | A | T10 | Activity based training | 140-210 | 50 \| 5-20mA (suprathreshold) \| 100-400 | Paddle | L1-S1 | hip/knee/ankle | none | Continuation of Wan et al., 2024 |
| Albano et al., 2025[49] | NCT05926843 | Europe | 1 | 33 | 29 | 4 | M | Lower limb | NR | C | T11-T12 | Activity based training | 240 | 30-40 \| subthreshold \| 300 | Paddle | L1-L3 | hip | none | First study investigating conus medullaris injuries |

Abbreviations: AIS – American Spinal Injury Association Impairment Scale; NCT – National Clinical Trial; NR – not reported; SCS – spinal cord stimulation. Values are presented as mean (SD) or median [range], unless otherwise noted. * Indicates that the study includes one or more previously reported patients; values reflect newly reported cases only.

**Supplementary Table 3. Summary of non-randomized and randomized clinical trials of transcutaneous SCS for motor recovery after spinal cord injury**

| Study information | | | Demographics | | | | | | SCI demographics | | | Rehabilitation | | Transcutaneous SCS | | | | Safety | |
|---|---|---|---|---|---|---|---|---|---|---|---|---|---|---|---|---|---|---|---|
| **Study Name** | **NCT** | **Study region** | **Sample size** | **Age at time of study (yrs)** | **Age at time of injury (yrs)** | **Time since injury (yrs)** | **Sex** | **Upper or Lower limb** | **Cause of injury** | **AIS grade** | **Neurological level of injury** | **Type of rehab** | **Hours on avg** | **Parameters: Carrier Frequency \| Frequency (Hz) \| Amplitude (V or mA) \| Pulse width (µs)** | **Lead location: Cathode \| Anode** | **Spinal levels targeted** | **Joint targeted** | **Adverse events** | **Notes** |
| Gad et al., 2018[37] | NCT01906424 | USA | 6** | 40.2 (16.6) | 30.2 (13.3) | 10.0 (7.1) | M: 5 F: 1 | Upper limb | NR | B: 2 C: 4 | C4: 3 C6: 2 C8: 1 | Activity based training | 8-16 | 10kHz carrier \| 30 \| 10-250mA (suprathreshold) \| 1000 | C3-4/C6-7 \| iliac crest | C3-4/C6-7 | hand | 1 (100%); UTI | 2 participants discontinued; 1 due to UTI, 1 due to educational |
| Inanici et al., 2018[32] | NCT03184792 | USA | 1 | 62 | 60 | 2 | M | Upper limb | Sports: 1 | D | C3 | Activity based training | 72-90 | 10kHz carrier \| 30 \| 80-120mA (subthreshold) \| 1000 | C3-4/C6-7 \| iliac crest | C3-4/C6-7 | hand | 1 (100%); Mild skin irritation, resolved <10min | |
| Estes et al., 2021[34] | NCT03240601 | USA | 8 | 43.6 (18.1) | 43.4 (18.1) | 0.3 (0.1) | M | Lower limb | NR | C: 2 D: 6 | C1: 1 C2: 1 C3: 1 C4: 2 C5: 2 C7: 1 | Activity based training | 72 | NR | T11/T12 \| abdomen | NR | hip/knee/ankle | 8 (100%); Mild discomfort during stimulation | |
| Inanici et al., 2021[33] | NCT03184792 | USA | 6 | 42.7 (14.2) | 38.1 (14.9) | 4.6 (3.8) | M: 4 F: 2 | Upper limb | NR | B: 2 C: 2 D: 2 | C3: 1 C5: 5 | Activity based training | 96 | 10kHz carrier \| 30 \| 40-90mA (subthreshold) \| 1000 | Above/below lesion \| iliac crest | NR | hand | 1 (100%); Mild allergic skin reaction | |
| Samejima et al., 2022[76] | NCT03509558 | USA | 2 | 64.0 (0.0) | 60.0 (0.7) | 4.0 (0.7) | M: 2 | Lower limb | NR | D: 2 | C4: 1 C6: 1 | Activity based training | 72-128 | 10kHz carrier \| 30 \| 5-40mA cervical; 35-75mA lumbar (subthreshold) \| 1000 | Cervical: C3-4/C6-7 \| iliac crest Lumbar: T11/L1 \| iliac crest | C3-4/C6-7; T11-L1 | hip/knee/ankle | none | |
| Chandrasekaran et al., 2023[77] | NCT04755699 | USA | 2 | 27.5 (10.6) | 22.0 (8.5) | 5.5 (2.1) | M: 2 | Upper limb | NR | A: 1 B: 1 | C5: 2 | Activity based training | 32 | 10kHz carrier \| 50 \| 140-160mA (suprathreshold) \| 500 | C1-C8 \| lumbar spine | C6 | Arm (triceps) | none | |
| García-Alén et al., 2023[56] | NCT07140354 | Europe | 15 | 37.4 (13.3) | 36.9 (13.3) | 0.5 (0.2) | M: 14 F: 1 | Upper limb | NR | A: 2 B: 3 C: 4 D: 6 | C3: 1 C4: 5 C5: 5 C6: 2 C7: 2 | Activity based training | 32-40 | 10kHz carrier \| 30 \| 39-86mA (subthreshold) \| 1000 | C3-4/C6-7 \| iliac crest | C3-4/C6-7 | hand | 15 (100%); Skin irritation | |
| Moritz et al., 2024[15] | NCT04697372 | Europe, USA | 60 | 47.2 (15.5) | 41.1 (16.2) | 6.1 (7.5) | M: 50 F: 10 | Upper limb | Sports: 17 Fall: 15 Recreation: 14 | B: 9 C: 28 D: 23 | C2: 9 C3: 3 C4: 16 C5: 13 C6: 13 | Activity based training | 25 | 10kHz carrier \| 30 \| 0-180mA (subthreshold) \| 100 | Above/below lesion \| iliac crest / clavicle. | C3-T1 | Hand | 17 (26.6%*) events related | |

| | | | | | | | | | | | | | | | | | | |
|---|---|---|---|---|---|---|---|---|---|---|---|---|---|---|---|---|---|---|
| | | | | | | | | | Vehicular: 12<br>Other: 2 | | C7: 6 | | | | Most common cathodes:<br>C3-4/C6-7 (~50%);<br>C4-5/C6-7 (~20%);<br>C3-4/C7-T1 (~10%) | | | to the device<br><br>*included safety population |
| Comino-Suarez et al., 2025[68] | NCT05210166 | Europe | 14 | 47.4 (14.5) | 46.4 (14.5) | 0.3 (0.1) | M: 14 | Lower limb | Traumatic: 7<br>Other: 8 | C: 8<br>D: 6 | C4: 6<br>C6: 1<br>T4: 1<br>T6: 1<br>T9: 1<br>T10: 4 | Activity based training | 20 | No carrier \| 20 \| subthreshold \| 1000 | T11-T12 \| umbilicus | L1-S1 | hip/knee/ankle | 14 (100%): Skin irritation; 12 (85.6%): discomfort during stimulation |
| Bye et. al., 2026[16] | ACTRN12620001241921 | Australia, Europe, USA | 25 | 46 [IQR: 37-56] | NR | > 1 year | M: 20<br>F: 5 | Lower limb | NR | A: 4<br>B: 0<br>C: 16<br>D: 5 | C7-T5: 11<br>T6-T11: 14 | Activity based training | 18 | 10kHz carrier \| 20 \| subthreshold \| 1000 | L1-2 \| abdomen | L1-S1 | hip/knee/ankle | 56 events; 22 (39.2%) skin irritation; 6 (10.7%) increase spasticity; 6 (10.7%) pain |

Abbreviations: AIS – American Spinal Injury Association Impairment Scale; NCT – National Clinical Trial; NR – not reported; SCS – spinal cord stimulation. Values are presented as mean (SD) or median [range], unless otherwise noted. * Indicates that the study includes one or more previously reported patients; values reflect newly reported cases only.

**Supplementary Table 4. Summary of cohort and case studies of epidural SCS for motor recovery after spinal cord injury**

| Study information | | Demographics | | | | | | SCI demographics | | | Rehabilitation | | Epidural SCS | | | | Safety | |
|---|---|---|---|---|---|---|---|---|---|---|---|---|---|---|---|---|---|---|
| **Study Name** | **Study region** | **Sample size** | **Age at time of study (yrs)** | **Age at time of injury (yrs)** | **Time since injury (yrs)** | **Sex** | **Upper or Lower limb** | **Cause of injury** | **AIS grade** | **Neurological level of injury** | **Type of rehab** | **Hours on avg** | **Parameters Frequency (Hz) \| Amplitude (V or mA) \| Pulse width (µs)** | **Type of lead** | **Spinal levels targeted** | **Joint targeted** | **Adverse events** | **Notes** |
| Dimitrivec, et al., 1998[26] | Europe/USA | 6 | 34.7 (14.1) | 31.0 (13.7) | 3.7 (2.3) | M: 3 F: 3 | Lower limb | Vehicular: 5 Fall: 1 | A: 6 | C5-6: 1 T3: 2 T4: 2 T7: 1 | NR | NR | 25-60 \| 5-9V (suprathreshold) \| NR | Perc | L1-S1 | hip/knee/ankle | none | |
| Herman et al., 2002[78] | USA | 1 | 43 | 39.5 | 3.5 | M | Lower limb | NR | C: 1 | C5-6: 1 | Partial weight bearing therapy | NR | 20-60 \| (suprathreshold) \| 800 | Perc | L1-S1 | hip/knee/ankle | none | Improved cardiovascular, respiratory, and metabolic effects |
| Jilge et al., 2004[47] | Europe | 5 | 27.6 (3.8) | 22.6 (5.5) | 5.0 (2.4) | M: 2 F: 3 | Lower limb | Vehicular: 4 Sport: 1 | A: 4 B: 1 | C4-5: 1 C5-6: 1 T4-5: 1 T5-6: 1 T7: 1 | NR | NR | 5-50 \| 5-10V (suprathreshold) \| 210 | Perc | L1-S1 | knee extension | none | |
| Minassian et al., 2004[79] | Europe | 10 | 26.9 (11.7) | 24.2 (11.8) | 2.7 (1.3) | M: 7 F: 3 | Lower limb | Vehicular: 8 Sport: 1 Fall: 1 | A: 8 B: 2 | C4: 2 C6: 1 C7: 1 T4: 2 T5: 1 T6: 1 T7: 1 T10: 1 | NR | NR | 25-50 \| 6-10V (suprathreshold) \| 210 | Perc | L1-S1 | hip/knee/ankle | none | |
| Huang et al., 2006 [*80] | USA | 1* | 48 | 40 | 8 | M | Lower limb | NR | C | T8 | Partial weight bearing therapy | NR | 20-40 \| <3.5V (subthreshold) \| 200-500 | Perc | T10-L2 | hip/knee | none | Continuation of Herman et al., 2002 |
| Harkema et al., 2011[11] | USA | 1 | 23.8 | 20.4 | 3.4 | M | Lower limb | Vehicular | B | T2 | Activity based training | 108 locomotor 54 standing | 5-40 \| 0.5-10V (suprathreshold) \| 210, 450 | Paddle | L1-S1 | hip/knee/ankle | none | Also gained bladder, sexual function, and improved temperature control |
| Angeli et al., 2014[50] | USA | 3* | 27.9 (4.5) | 25.0 (3.4) | 2.9 (1.1) | M: 3 | Lower limb | Vehicular: 3 | A: 2 B: 1 | C7: 1 T4: 1 T5: 1 | Activity based training | 80 | 5-40 \| 0.5-7.5V (suprathreshold) \| 450 | Paddle | L1-S1 | hip/knee/ankle | none | Continuation of Harkema et al., 2011 |
| Rejc et al., 2015 [*81] | USA | * | - | - | - | - | - | - | - | - | - | - | - | - | - | - | none | Same participants as Angeli et al., 2014 |
| Lu et al., 2016[44] | USA | 2 | 19.0 (1.4) | 16.5 (0.7) | 2.3 (0.4) | M: 2 | Upper limb | Sports: 2 | B: 2 | C5: 1 C6: 1 | Exp* | >60 | 2-40 \| 0.1-10mA \| 210 | Paddle: 1 Perc: 1 | C4-T1 | arm/hand | none | First in-human evidence for cervical SCS |
| Grahn et al., 2017[82] | USA | 1 | 26 | 23 | 3 | M | Lower limb | Trauma | A | T6 | Activity based training | 40-56 | 28-44 \| 2.9-16 (suprathreshold) \| 400-500 | Paddle | L1-S1 | hip/knee/ankle | None | |
| Rejc et al., 2017 [*83] | USA | * | - | - | - | - | - | - | - | - | - | - | - | - | - | - | none | Same participants as Angeli et al., 2014; Rejc et al., 2015 |
| Rejc et al., 2017 [*84] | USA | * | - | - | - | - | - | - | - | - | - | - | - | - | - | - | none | Same participant as |

| | | | | | | | | | | | | | | | | | | |
|---|---|---|---|---|---|---|---|---|---|---|---|---|---|---|---|---|---|---|
| | | | | | | | | | | | | | | | | | | Angeli et al., 2014; Rejc et al., 2015; Rejc et al., 2017 |
| Gill et al., 2018 *[85] | USA | * | - | - | - | - | - | - | - | - | - | - | - | - | - | - | none | Same participant as Grahn et al., 2017; participant can now walk independently only with eSCS ON |
| Gorgey et al., 2020[86] | USA | 1 | 34 | 30 | 4 | M | Lower limb | Sports | A | C7 | Exoskeleton assisted walking | NR | 40 \| 4.4-8V (suprathreshold) \| 420µs | Paddle | T12-S2 | hip/knee/ankle | none | |
| Krucoff et al., 2020[87] | USA | 1 | 48 | 41 | 7 | M | Lower limb | Trauma | A | L1 | Physical therapy | 1560-2340 | 40, 60 \| 2-11.5mA (suprathreshold) \| 250µs | Paddle | L1-S1 | hip/knee/ankle | none | Bowel incontinence improved from 3-4 episodes per week to 2-3 per month |
| Mesbah et al., 2021[88] | USA | 20 | 31.0 (9.9) | 24.7 (8.6) | 6.3 (3.5) | M: 15 F: 5 | Lower limb | NR | A: 14 B: 6 | C3: 1 C4: 9 C5: 4 C6: 2 C8: 1 T1: 1 T3: 1 T4: 1 | NR | NR | 25-30 \| suprathreshold \| 450-1000µs | Paddle | L2-S2 | hip/knee/ankle | none | |
| Gorgey et al., 2022[89] | USA | 1 | 25 | 21.33 | 3.67 | M | Lower limb | NR | A | T3 | NR | NR | 20-40 \| 5.5-6 (suprathreshold) \| 210-240µs | Perc | L1-S2 | hip/knee/ankle | 1; Lead migration | The left lead migrated completely out of the epidural space. Right lead migrated down 1-2 levels |
| Kandhari et al., 2022[53] | India | 10 | 32.9 (8.9) | 29.1 (9.7) | 3.8 (4.5) | M: 9 F: 1 | Lower limb | NR | A: 10 | T2: 1 T5: 2 T6: 2 T8: 1 T9: 1 T10: 1 T11: 1 T12: 1 | Activity based training | 72 | 15-60 \| 1-1.5V (subthreshold) \| 210-400 | Paddle | T12-S2 | hip/knee/ankle | none | |
| Angeli et al., 2024 *[59] | USA | * | - | - | - | - | - | - | - | - | - | - | - | - | - | - | none | Continuation of participants in Angeli et al., 2014, Rejc et al., 2017, Mesbah et al., 2019; describes improvements in bladder, cardiovascular, and motor function |

Abbreviations: AIS – American Spinal Injury Association Impairment Scale; NCT – National Clinical Trial; NR – not reported; SCS – spinal cord stimulation. Values are presented as mean (SD) or median [range], unless otherwise noted. * Indicates that the study includes one or more previously reported patients; values reflect newly reported cases only.

**Supplementary Table 5. Summary of cohort and case studies of transcutaneous SCS for motor recovery after spinal cord injury**

| Study information | | Demographics | | | | | | SCI demographics | | | Rehabilitation | | Transcutaneous SCS | | | | Safety | |
|---|---|---|---|---|---|---|---|---|---|---|---|---|---|---|---|---|---|---|
| **Study Name** | **Study region** | **Sample size** | **Age at time of study (yrs)** | **Age at time of injury (yrs)** | **Time since injury (yrs)** | **Sex** | **Upper or Lower limb** | **Cause of injury** | **AIS grade** | **Neurological level of injury** | **Type of rehab** | **Hours on avg** | **Parameters: Carrier Frequency \| Frequency (Hz) \| Amplitude (V or mA) \| Pulse width (µs)** | **Lead location: Cathode \| Anode** | **Spinal levels targeted** | **Joint targeted** | **Adverse events** | **Notes** |
| Gerasimenko et al., 2015[39] | USA | 5 | 31.4 (16.8) | 28.2 (15.7) | 3.2 (1.6) | M: 5 | Lower limb | Sports: 4 Vehicular: 1 | B: 5 | C5-6: 1 C6:1 C7: 1 T3-4: 2 | NR | NR | 10kHz carrier \| 30 (T11), 5 (Co1) \| 80-180mA (suprathreshold) \| 1000 | T11, Co1, or T11/Co1 \| iliac crests | T11/Co1 | hip/knee/ankle | none | |
| Hofstoetter et al., 2020[90] | Europe | 12 | 41.3 (19.1) | 29.2 (15.5) | 12.1 (16.2) | M: 9 F: 3 | Lower limb | Vehicular: 4 Sport: 1 | A: 3 C: 3 D: 6 | C4: 3 C6: 1 C7: 3 T4: 2 T6: 2 T7: 1 | NR | 0.5: avg 15 in one participant | no carrier \| 50 \| 16-100mA (subthreshold) \| 1000 | T11-T12 \| abdomen | L1-S1 | hip/knee/ankle | 1; UTI | |
| Zhang et al., 2020[91] | USA | 1 | 38 | 23 | 15 | M | Upper limb | Sports: 1 | A | C5 | Task-specific training | 18 | 10kHz carrier \| 30 \| 30-60mA (C3/4), 10-20mA (C7/T1) \| (subthreshold) \| 1000 | C3-4/C7-T1 \| iliac crests | C3-4/C7-T1 | hand | none | |
| McHugh et al., 2020[92] | USA | 10 | 43.0 (17.7) | 29.6 (16.7) | 13.4 (16.7) | M: 6 F: 4 | Lower limb | Non-Trauma: 6 Trauma: 4 | C: 4 D: 6 | C4: 1 C5: 2 C6: 1 C7: 1 T1: 1 T3: 1 T4: 1 T8: 1 T9: 1 | Walking based therapy | 48 | no carrier \| 50 \| 20-80mA (subthreshold) \| 1000 | T11-T12 \| abdomen | L1-S1 | hip/knee/ankle | 4; mild skin reaction and mild incontinence after stimulation | |
| Meyer et al., 2020[93] | Europe | 10 | 45.4 (12.4) | 33.8 (13.8) | 11.6 (10.2) | M: 9 F: 1 | Lower limb | NR | D: 10 | C3: 1 C4: 1 C5: 2 C6: 1 C7: 2 T3: 1 T4: 1 T10: 1 | Activity based training | NR | no carrier \| 30 \| 15-70mA (subthreshold) \| 1000 | T11-T12 \| abdomen | L2-S2 | ankle | none | |
| Siu et al., 2022[94] | USA | 1 | 27 | 22 | 5 | M | Lower limb | Trauma | A | T2 | Activity based training | 60 | 5kHz carrier \| 30-40 \| 2.5mA (subthreshold) \| 1000 | C3-C4, T11-T12, L2-L3, sacral-coccygeal \| clavicle, abdomen iliac crest | C3-C4, T11-T12, L2-L3, S3-4 | hip/knee/ankle | none | |
| Sharma et al., 2023[95] | USA | 1 | 53 | 45 | 8 | M | Upper limb | Sports | A | C2 | Activity based training | 60 | 5kHz carrier \| 30-40 \| 2.5mA (subthreshold) \| 1000 | C3-4, C4-5, C6-7, C7-T1, T1-2, T11-T12 \| clavicle | C3-T1 | elbow/wrist/hand | 1; Blistering and skin reactions | Multi-site tSCS was superior to single site |

| | | | | | | | | | | | | | | | | | |
|---|---|---|---|---|---|---|---|---|---|---|---|---|---|---|---|---|---|
| Verma et al., 2025[96] | USA | 5 | 41.5 (19.6) | 32.5 (18.2) | 9.0 (5.9) | M: 4 F: 1 | Upper limb | NR | A: 3 B: 1 C: 1 | C4-5: 2 C5: 1 C5-6: 1 C6-7: 1 | NR | NR | No carrier \| 30 \| 55-75mA (subthreshold) \| 500 | C3-4/C6-7 \| iliac crests | C3-T1 | hand | none |

Abbreviations: AIS – American Spinal Injury Association Impairment Scale; NCT – National Clinical Trial; NR – not reported; SCS – spinal cord stimulation. Values are presented as mean (SD) or median [range], unless otherwise noted.

**Supplementary Table 6. Participant demographics and SCS parameters stratified by studies targeting upper limb (cervical SCS) or lower limb (lumbar SCS)**

| | **Overall** | | **Upper Limb / Cervical SCS** | | **Lower Limb / Lumbar SCS** | |
|---|---|---|---|---|---|---|
| | **Epidural SCS** | **Transcutaneous SCS** | **Epidural SCS** | **Transcutaneous SCS** | **Epidural SCS** | **Transcutaneous SCS** |
| **Total Number of Participants** | 94 | 184 | 2 | 97 | 92 | 87 |
| **Age at time of study (yrs)** | 32.1 ± 10.3 | 42.1 ± 15.9 | 19.0 ± 1.4 | 39.7 ± 14.6 | 32.5 ± 10.3 | 43.6 ± 16.5 |
| **Age at time of injury (yrs)** | 27.4 ± 9.9 | 35.8 ± 16.0 | 16.8 ± 1.1 | 34.9 ± 14.4 | 27.6 ± 9.8 | 36.5 ± 16.9 |
| **Time since injury (yrs)** | 4.8 ± 3.3 | 6.1 ± 9.9 | 2.3 ± 0.4 | 4.8 ± 5.6 | 4.9 ± 3.3 | 6.9 ± 11.7 |
| **Sex (M/F)** | **M:** 74 (78.7)<br>**F:** 20 (21.3) | **M:** 154 (83.7)<br>**F:** 30 (16.3) | **M:** 2 (100.0)<br>**F:** 0 (0.0) | **M:** 82 (84.5)<br>**F:** 15 (15.5) | **M:** 72 (78.2)<br>**F:** 20 (21.7) | **M:** 72 (82.8)<br>**F:** 15 (17.2) |
| **AIS Status** | **A:** 68 (72.3)<br>**B:** 18 (19.1)<br>**C:** 7 (7.5)<br>**D:** 1 (1.1) | **A:** 16 (8.7)<br>**B:** 23 (12.5)<br>**C:** 72 (39.1)<br>**D:** 73 (39.7) | **A:** 0 (0.0)<br>**B:** 2 (100.0)<br>**C:** 0 (0.0)<br>**D:** 0 (0.0) | **A:** 8 (8.2)<br>**B:** 18 (18.6)<br>**C:** 39 (40.2)<br>**D:** 32 (33.0) | **A:** 68 (74.0)<br>**B:** 16 (17.4)<br>**C:** 7 (7.6)<br>**D:** 1 (1.0) | **A:** 8 (9.2)<br>**B:** 5 (5.7)<br>**C:** 33 (37.9)<br>**D:** 41 (47.1) |
| **Neurological Level of Injury*** | **C1-C8:** 34 (36.2)<br>**T1-T5:** 36 (38.3)<br>**T5-below:** 24 (25.5) | **C1-C8:** 136 (78.6)<br>**T1-T5:** 11 (6.4)<br>**T5-below:** 26 (15.0) | **C1-C8:** 2 (100.0) | **C1-C8:** 97 (100.0) | **C1-C8:** 32 (34.8)<br>**T1-T5:** 36 (39.1)<br>**T5-below:** 24 (26.1) | **C1-C8:** 39 (51.3)<br>**T1-T5:** 11 (14.5)<br>**T5-below:** 26 (34.2) |
| **SCS Parameters Frequency (Hz)** | **0-10:** 1.9%<br>**11-20:** 6.9%<br>**21-30:** 44.6%<br>**31-40:** 22.7%<br>**41-50:** 13.6%<br>**51-60:** 5.3%<br>**61-100:** 4.3%<br>**100+:** 0.8% | **0-10:** 1.4%<br>**11-20:** 14.2%<br>**21-30:** 70.5%<br>**31-40:** 0.3%<br>**41-50:** 13.6% | **0-10:** 25%<br>**11-20:** 25%<br>**21-30:** 25%<br>**31-40:** 25%<br>**41-50:** 0%<br>**51-60:** 0%<br>**61-100:** 0%<br>**100+:** 0% | **0-10:** 0%<br>**11-20:** 0%<br>**21-30:** 97.9%<br>**31-40:** 0%<br>**41-50:** 2.1% | **0-10:** 1.4%<br>**11-20:** 6.5%<br>**21-30:** 45.0%<br>**31-40:** 22.7%<br>**41-50:** 13.9%<br>**51-60:** 5.4%<br>**61-100:** 4.4%<br>**100+:** 0.8% | **0-10:** 3.2%<br>**11-20:** 31.6%<br>**21-30:** 37.3%<br>**31-40:** 1.3%<br>**41-50:** 26.6% |
| **Amplitude** | **Suprathreshold:** 83.7%<br>**Subthreshold:** 16.3% | **Suprathreshold:** 7.4%<br>**Subthreshold:** 92.6% | **Suprathreshold:** NA<br>**Subthreshold:** NA | **Suprathreshold:** 7.2%<br>**Subthreshold:** 92.8% | **Suprathreshold:** 83.7%<br>**Subthreshold:** 16.3% | **Suprathreshold:** 6.3%<br>**Subthreshold:** 93.7% |
| **Pulse width (μs)** | **0-200:** 2.7%<br>**201-250:** 30.6%<br>**251-300:** 11.2%<br>**301-350:** 5.3%<br>**351-400:** 5.3%<br>**401-450:** 28.6%<br>**451-500:** 2.2%<br>**800:** 2.4%<br>**1000:** 11.8% | **100:** 34.1%<br>**500:** 4.0%<br>**1000:** 61.9% | **210:** 100% | **100:** 61.9%<br>**500:** 7.2%<br>**1000:** 30.9% | **0-200:** 2.8%<br>**201-250:** 28.9%<br>**251-300:** 11.5%<br>**301-350:** 5.4%<br>**351-400:** 5.4%<br>**401-450:** 29.3%<br>**451-500:** 2.2%<br>**800:** 2.4%<br>**1000:** 12.0% | **1000:** 100% |

Abbreviations: AIS – American Spinal Injury Association Impairment Scale; SCS – spinal cord stimulation; Continuous values are presented as mean ± SD. Categorical values are presented as count (percent). SCS parameters are presented as a percentage of all reported instances. Highlighted cells represent the highest frequency instance within each category. *This only represents the studies that describe the neurological level of injury explicitly.

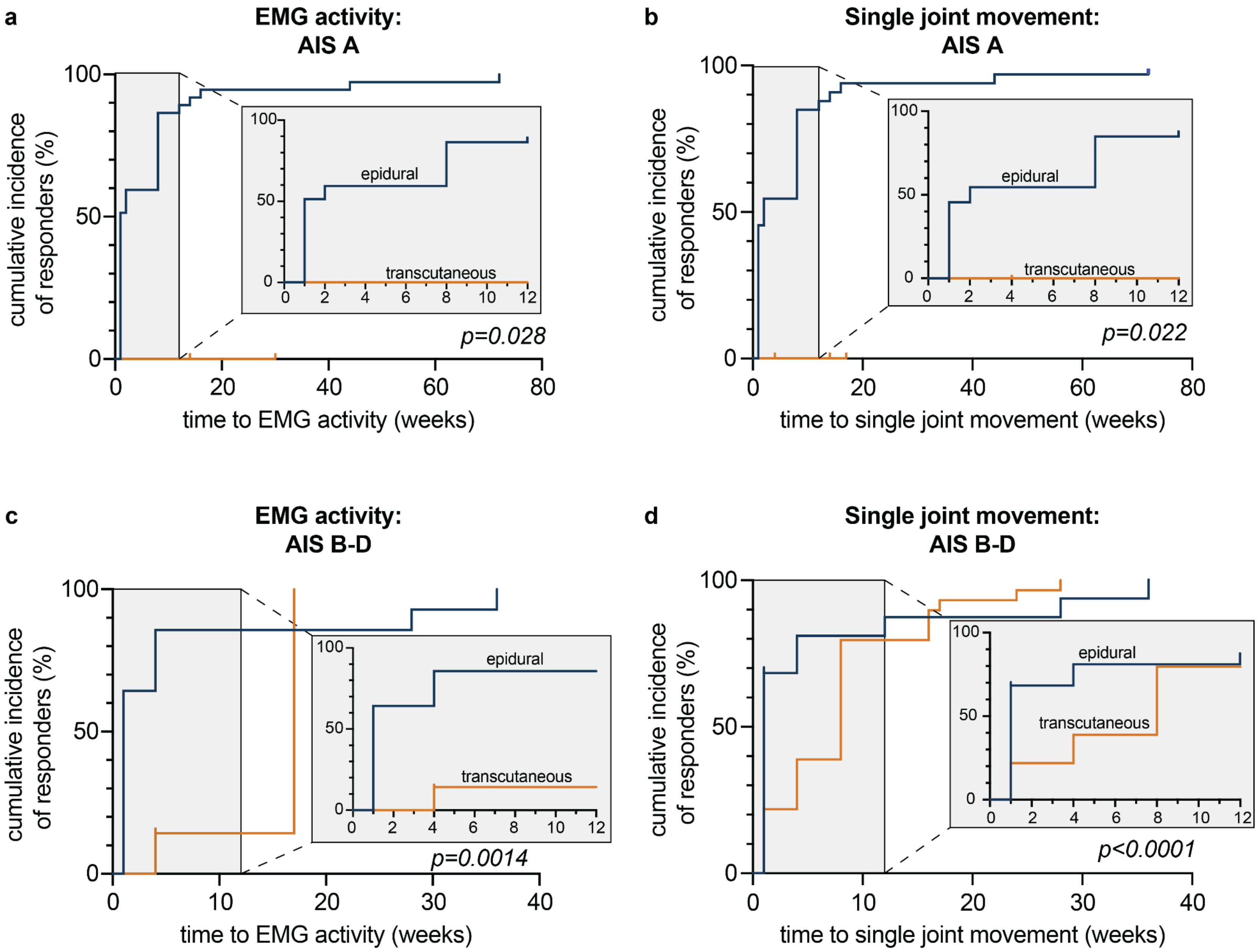
a
EMG activity:
AIS A
cumulative incidence
of responders (%)
time to EMG activity (weeks)
epidural
transcutaneous
p=0.028
b
Single joint movement:
AIS A
cumulative incidence
of responders (%)
time to single joint movement (weeks)
epidural
transcutaneous
p=0.022
c
EMG activity:
AIS B-D
cumulative incidence
of responders (%)
time to EMG activity (weeks)
epidural
transcutaneous
p=0.0014
d
Single joint movement:
AIS B-D
cumulative incidence
of responders (%)
time to single joint movement (weeks)
epidural
transcutaneous
p<0.0001

**Supplementary Fig. 1 | Determinants of response to epidural and transcutaneous SCS stratified by injury severity.**
**a.** Cumulative time to recovery of volitional EMG activity in participants with sensorimotor complete injuries (AIS A), showing that eSCS cohorts achieve recovery significantly faster than tSCS cohorts ($p=0.028$). **b.** Cumulative time to recovery of volitional single joint movement in participants with AIS A injuries, demonstrating a similar pattern faster response in eSCS compared to tSCS cohorts ($p=0.022$). **c.** Cumulative time to recovery of volitional EMG activity in participants with sensory incomplete injuries (AIS B-D), showing that eSCS cohorts achieve most recovery faster, while tSCS achieves meaningful cumulative response rates over longer treatment durations ($p=0.0014$). **d.** Cumulative time to recovery of volitional single joint movement in participants with AIS B-D injuries, demonstrating that eSCS cohorts achieve faster rates of recovery ($p<0.0001$). Insets show an expanded view of the first 12 weeks of follow-up.

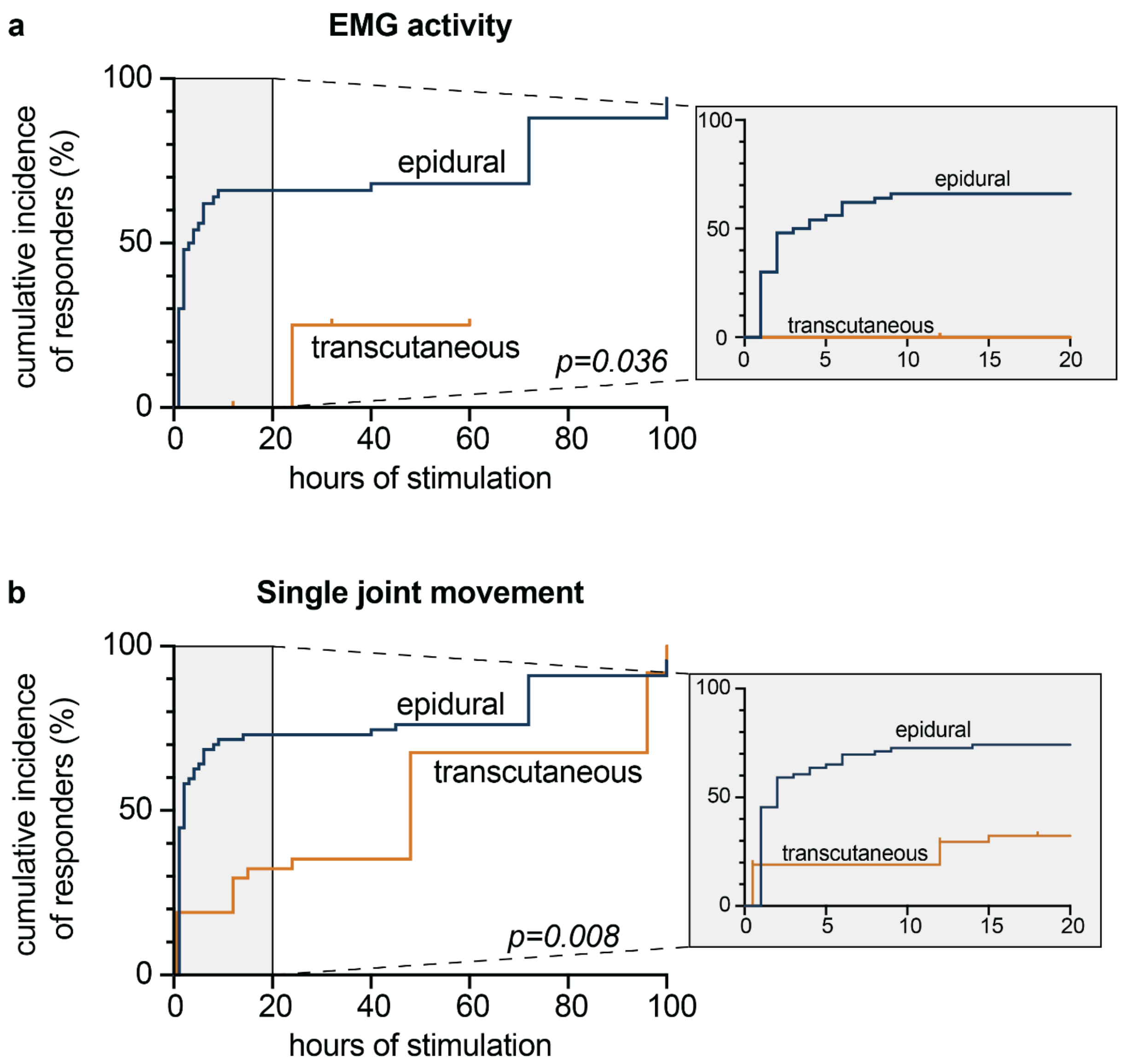
a
EMG activity
cumulative incidence of responders (%)
100
50
0
epidural
transcutaneous
p=0.036
0
20
40
60
80
100
hours of stimulation
100
50
0
epidural
transcutaneous
0
5
10
15
20
b
Single joint movement
cumulative incidence of responders (%)
100
50
0
epidural
transcutaneous
p=0.008
0
20
40
60
80
100
hours of stimulation
100
50
0
epidural
transcutaneous
0
5
10
15
20

**Supplementary Fig. 2 | Determinants of response to epidural and transcutaneous SCS controlling for stimulation dose.**
**a.** Cumulative incidence of responders achieving recovery of volitional EMG activity as a function of total hours of stimulation delivered, demonstrating that eSCS achieves significantly faster response compared to tSCS even when controlling for stimulation dose ($p=0.036$). **b.** Cumulative incidence of responders achieving recovery of volitional single joint movement as a function of total hours of stimulation delivered, showing that eSCS achieves response after a fewer number of hours of stimulation relative to tSCS after accounting for stimulation dose ($p=0.008$). Insets show an expanded view of the first 20 hours of stimulation.
